# A Proteogenomic Signature of Age-related Macular Degeneration in Blood

**DOI:** 10.1101/2021.07.27.21261194

**Authors:** Valur Emilsson, Elias F. Gudmundsson, Thorarinn Jonmundsson, Michael Twarog, Valborg Gudmundsdottir, Nancy Finkel, Stephen Poor, Xin Liu, Robert Esterberg, Yiyun Zhang, Sandra Jose, Chia-Ling Huang, Sha-Mei Liao, Joseph Loureiro, Qin Zhang, Cynthia L Grosskreutz, Andrew A Nguyen, Qian Huang, Barrett Leehy, Rebecca Pitts, Brynjolfur G. Jonsson, Thor Aspelund, John R. Lamb, Fridbert Jonasson, Lenore J. Launer, Mary Frances Cotch, Lori L. Jennings, Vilmundur Gudnason, Tony E. Walshe

## Abstract

Age-related macular degeneration (AMD) is one of the most frequent causes of visual impairment in the elderly population. The overall etiology of AMD is complex and still poorly understood, though age, obesity, smoking, and high-density lipoprotein are known risk factors. In one of the first successful reported genome-wide association studies (GWAS), common genetic variants were strongly associated with AMD, including variants within the complement factor H (CFH) gene. To date, 34 genomic regions have been linked to AMD; however, the genes that mediate the risk remain largely unknown, indicating that novel approaches to identifying causal candidates are needed. Recent advances in proteomic technology have exposed the serum proteome’s depth and complexity. In the Age, Gene/Environment Susceptibility Reykjavik Study (AGES-RS), a broad population-based study of the elderly (N = 5764), levels of 4137 human serum proteins and associated networks were integrated with established genetic risk loci for AMD, revealing many predicted as well as novel proteins and pathways, linked to the disease. Serum proteins were also found to reflect AMD severity independent of genetics and predict progression from early to advanced AMD after five years in this population. A two-sample Mendelian randomization study of five proteins associated with AMD found CFHR1, CFHR5, and FUT5 to be causally related to the disease, all of which were directionally consistent with the observational estimates. This study provides a robust and unique framework for elucidating the pathobiology of AMD.

## Introduction

AMD is a progressive late-onset disease that primarily affects the macular area of the retina and is a common cause of permanent loss of vision in the elderly population^1^. The clinical hallmark of early AMD is an accumulation of extracellular protein and lipid containing deposits between the retinal pigment epithelium (RPE) and Bruch’s membrane, termed drusen. Advanced AMD can be either neovascular (nAMD) associated with blood vessel growth and leakage, or geographic atrophy (GA/Dry AMD) characterized by patches of retinal pigment epithelial (RPE) cell and photoreceptor cell loss in the macula. Anti-VEGF therapy is highly effective in controlling the abnormal vessel growth and leakage in nAMD; however, the disease nevertheless progresses and by 7 years, 98% of nAMD patients have atrophy^2^. Patients with GA have marked visual disability with relentless visual deterioration and progression to legal blindness. There are currently no approved therapies for GA or early AMD^3,4^. Elucidation of AMD pathobiology and identification of modifiable targets are critical for identifying clinically relevant biomarkers and designing therapeutics.

The first common genetic risk factor reported for AMD was the missense variant rs1061170 in *CFH* at chromosome 1q31.3^5-8^. Many genes encoding other proteins involved in the complement cascade reside within the 1q31.3 region, including complement factors H related to 1 to 5 (*CFHR1-5*). Two independent variants at 1q31.3, rs1061170 and intronic variant rs1410996, account for 17% of AMD risk^9^. In the most recent GWAS examining 16,144 patients with advanced AMD, 52 independent variants were found at 34 different genomic loci explaining 46.7% of the variability in AMD risk^10^. The risk variants with the largest difference between late AMD patients and healthy controls reside within the *ARMS2/HTRA1* and *CFH* genomic loci, although for most variants, the effect size was small^10^. While some known AMD causal candidates are found at these genomic risk locations, the vast majority are yet to be identified.

Proteins are undeniably the key players in all life processes, with changes in their function and/or regulation influencing disease and well-being. As a result, changes in protein regulation and function, as well as their related networks, are most likely to mediate the genetic risk of complex diseases^11,12^. Serum proteins have the desired attributes required for a comprehensive and unified approach to measuring an individual’s global molecular status, as they capture information across many tissues and show a direct link to disease-related molecular pathways and activities^11,12^.

Furthermore, serum proteins participate in cross-tissue regulatory loops, and therefore tissue specific disease progression emerges from an integration of local and systemic signals^12^. Recent developments in high-throughput measurements of thousands of proteins in a single sample have aided this work^11,13-15^, and aptamer-based affinity methods in particular, have been a driver of recent discoveries^11,16-19^. In this study, the serum levels of 4137 proteins measured in 5457 individuals from the prospective population-based AGES-RS were examined for association to different stages of AMD, disease progression, and the extent to which they mediated the genetics of the disease.

## Results

### Analysis of 4137 proteins in circulation found 28 proteins associated with various stages of AMD

Descriptive statistics on the data linked to AMD in the 5457 person AGES-RS cohort ages 67 years and older (mean age 76.6±5.6 years; 57.3% female) are shown in Supplementary Table S1. Using sex- and age-adjusted logistic regression analysis and two distinct definitions of early-stage AMD (Methods), we discovered that 28 serum proteins were associated with different stages of AMD using a study-wide significance threshold (Table 1, Figure 1a-c, Supplementary Figure S1a-c, and Supplementary Tables S2-3). Some of these proteins were only related to AMD’s early or late stages, like CEBPB and CFHR5, respectively (Figure 1d, e), while proteins such as FUT5 was associated with all stages of AMD (Figure 1f). We further stratified or combined late AMD into GA or nAMD and compared protein quintiles of all AMD-associated proteins with the various AMD-related outcomes. Levels of some AMD-associated proteins, such as CFHR1, BPIFB1, and CFHR5, increased almost continuously from no AMD to advanced nAMD (Supplementary Figure S2a-c). Supplementary Figure S3 depicts the distribution and relationship between the different quintiles of the 28 AMD-associated protein levels and the various AMD-related outcomes. In Supplementary Figure S4, the serum protein levels at the extreme end of the distribution, that is between the 5th and 1st quintiles using the first quintile as a reference (Methods), are shown in relation to the different AMD related outcomes. Again, it is evident that while some proteins were associated with all AMD stages (e.g., CFHR1, NDUFS4), some were more, or only associated with early (e.g., LINGO1, RAB17) or late-stage AMD (e.g., BIRC2) (Supplementary Figures S3 and S4). Finally, using single point sex and age-adjusted logistic regression analysis, we examined which if any of the 4137 proteins anticipated advancement to late AMD (pure GA or nAMD) while still in early AMD in the same people over a 5-year follow-up period. Only a single protein, PRMT3, showed significantly (OR = 1.88, P = 5.3×10^−6^) increased levels in early AMD at the baseline exam and prior to progression to pure GA at follow-up, after adjusting for multiple comparisons (Figure 1g). No baseline protein changes predicted advancement of early AMD to nAMD between baseline and five-year follow-up.

**Table 1:** Serum proteins significantly associated with prevalent AMD using regression analysis.

| Protein | Definition* | AMD any** |  |  | AMD early** |  |  | AMD late** |  |  |
| --- | --- | --- | --- | --- | --- | --- | --- | --- | --- | --- |
| | | β-value | P-value | P-corr | β-value | P-value | P-corr | β-value | P-value | P-corr |
| NDUFS4 | I, II | -0.298 | 1.4E-17 | 7.3E-14 | -0.264 | 2.8E-12 | 1.4E-08 | -0.562 | 2.7E-14 | 1.4E-10 |
| CFHR1 | I, II | 0.495 | 7.8E-10 | 3.9E-06 | 0.157 | 3.7E-06 | 0.01869 | 0.495 | 7.8E-10 | 3.9E-06 |
| FUT5 | I, II | 0.180 | 3.2E-08 | 0.00016 | 0.160 | 1.7E-06 | 0.00864 | 0.347 | 3.2E-06 | 0.01608 |
| TST | I, II | -0.235 | 2.6E-11 | 1.3E-07 | -0.218 | 8.1E-09 | 4.1E-06 | -0.353 | 1.2E-06 | 0.00629 |
| BAMBI | I | 0.177 | 5.0E-07 | 0.00255 | 0.158 | 8.2E-07 | 0.00413 | 0.138 | 0.06560 | ns |
| BPIFB1 | I, II | 0.193 | 3.1E-08 | 0.00016 | 0.176 | 1.9E-06 | 0.00980 | 0.271 | 0.00022 | ns |
| CEBPB | I | 0.173 | 9.7E-07 | 0.00489 | 0.171 | 2.6E-06 | 0.01432 | 0.189 | 0.01393 | ns |
| ST6GALNAC1 | I | 0.161 | 1.1E-06 | 0.00591 | 0.167 | 1.1E-06 | 0.00537 | 0.102 | ns | ns |
| DLL3 | I, II | 0.193 | 1.2E-07 | 0.00062 | 0.196 | 7.0E-07 | 0.00355 | 0.170 | 0.01847 | ns |
| UTS2 | I | 0.158 | 4.8E-06 | 0.02449 | 0.157 | 1.3E-05 | ns | 0.157 | 0.03319 | ns |
| TNFRSF14 | I | 0.147 | 5.3E-06 | 0.02696 | 0.158 | 2.2E-06 | 0.01129 | 0.123 | ns | ns |
| CXCL17 | I | 0.146 | 5.3E-06 | 0.02700 | 0.145 | 1.2E-05 | ns | 0.138 | 0.03687 | ns |
| RGS8 | I | 0.149 | 5.4E-06 | 0.02762 | 0.153 | 7.4E-06 | 0.03761 | 0.137 | 0.04946 | ns |
| RAB17 | I | 0.152 | 7.1E-06 | 0.03613 | 0.147 | 2.5E-05 | ns | 0.186 | 0.01027 | ns |
| HNRNPC | I | 0.143 | 7.4E-06 | 0.03769 | 0.137 | 3.0E-05 | ns | 0.163 | 0.01434 | ns |
| CRABP1 | I | 0.156 | 8.7E-06 | 0.04415 | 0.159 | 9.5E-06 | 0.04829 | 0.148 | ns | ns |
| CFHR5 | I, II | 0.397 | 3.2E-07 | 0.00160 | 0.077 | 0.01843 | ns | 0.397 | 3.2E-07 | 0.00160 |
| RPS6KB1 | II | 0.177 | 1.1E-06 | 0.00538 | 0.172 | 7.8E-06 | 0.03962 | 0.155 | 0.02414 | ns |
| ZBPB | II | 0.178 | 8.0E-06 | 0.04075 | 0.155 | 2.6E-05 | ns | 0.171 | 0.02920 | ns |
| KREMEN2 | II | 0.151 | 3.2E-05 | ns | 0.188 | 7.3E-06 | 0.03729 | 0.168 | 0.03047 | ns |
| CCL1 | I | 0.142 | 1.8E-05 | ns | 0.154 | 7.4E-06 | 0.03761 | 0.177 | 0.01066 | ns |
| CFP | II | -0.158 | 1.07E-05 | 0.04427 | -0.103 | 0.00079 | ns | -0.043 | ns | ns |
| GHR | II | -0.156 | 1.06E-05 | 0.04343 | -0.072 | 0.02341 | ns | -0.145 | 0.02935 | ns |
| LINGO1 | II | 0.162 | 8.0E-06 | 0.04065 | 0.120 | 0.00051 | ns | 0.172 | 0.01456 | ns |
| B3GNT6 | I | 0.142 | 1.0E-05 | 0.04302 | 0.104 | 7.2E-04 | ns | 0.121 | ns | ns |
| C2orf40 | I | 0.142 | 1.18E-05 | 0.04882 | 0.118 | 1.5E-04 | ns | 0.075 | ns | ns |
| BCL2L1 | I | 0.147 | 1.16E-05 | 0.04782 | 0.127 | 4.9E-05 | ns | 0.049 | ns | ns |
| BIRC2 | I | 0.107 | 0.00057 | ns | 0.086 | 0.00693 | ns | 0.321 | 5.0E-06 | 0.02635 |

\*Medical definition of early stage AMD by either Holliday et al. (PMID: 23326517), definition I, or by Jonasson et al. (PMID: 24768241) as definition II, where protein was significantly associated with AMD any, early and/or late.
\*\*If the association is found to be significant in both medical definitions of AMD (I and II), it is shown with the numerically lower P-value.
ns, not significant

**Figure 1.**
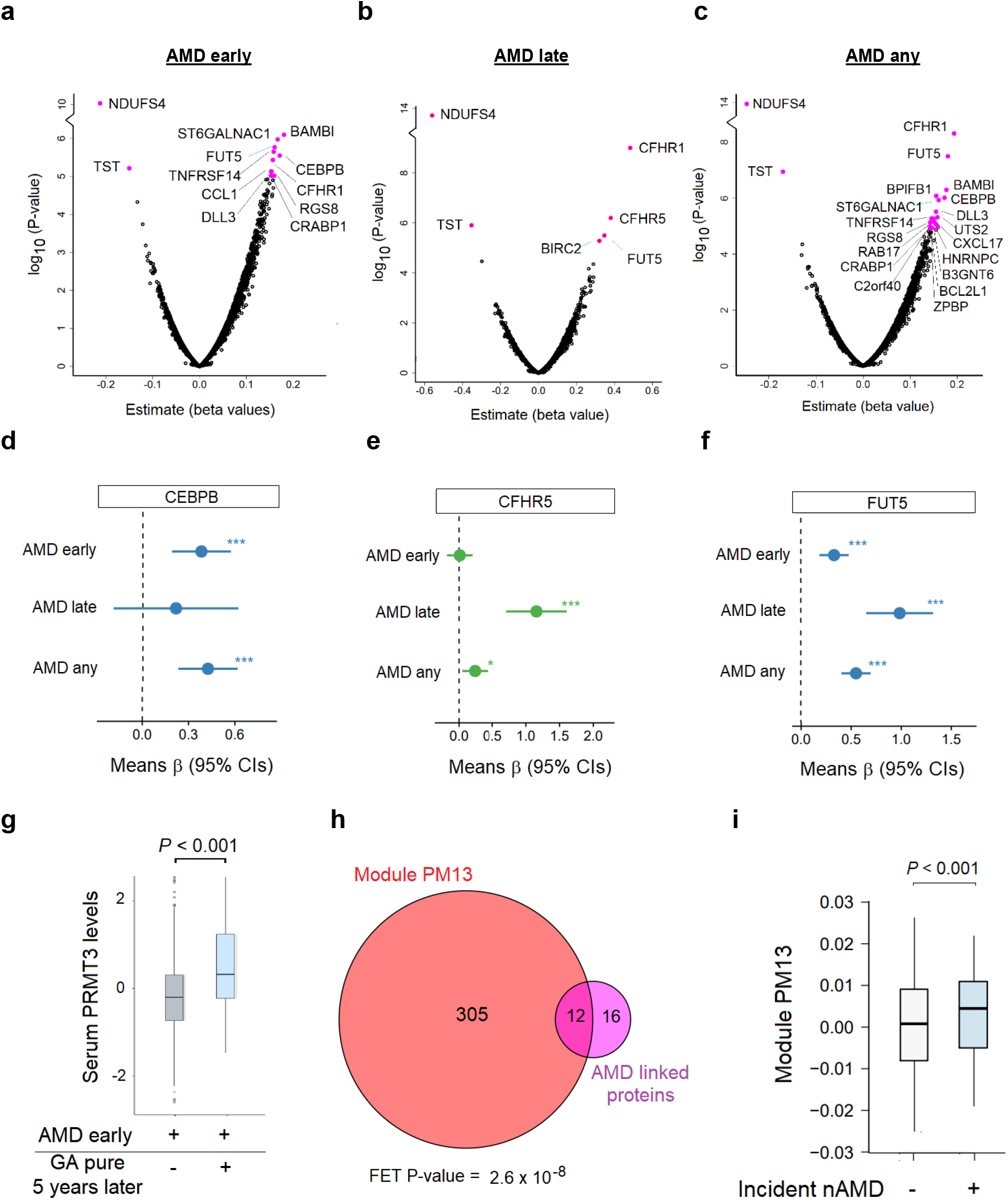
Association of global serum proteins with different stages of AMD. **a**. Using logistic regression analysis and Bonferroni correction for multiple comparisons, a volcano plot of all serum proteins associated with AMD early is shown, with colored data points highlighting significant associations. Definition of early-stage AMD was according to Holliday et al.^75^. **b**. A volcano plot of late stage AMD. **c**. Similar analysis and representive volcano plot for AMD any, where early-stage of the disease was defined according to Holliday et al.^75^. **d**. Relationship between the top and bottom quintiles for serum CEBPB levels and AMD-related outcomes. ***(P-value < 0.001), ** (P-value < 0.01), * (P-value < 0.05). **e**. Similar relationship shown for CFHR5, *** (P-value < 0.001), ** (P-value < 0.01), * (P-value < 0.05) and **f**. FUT5, *** (P-value < 0.001), ** (P-value < 0.01), * (P-value < 0.05). **g**. Raised PRMT3 levels in those with early AMD and who develop pure geographic atrophy (GA/Dry AMD) five years later (P-value = 5.3×10^−6^). **h**. Venn diagram showing significant enrichment (12 out of 28 proteins, P-value = 2.6×10^−8^) of the AMD-associated proteins in the previously described serum protein module PM13^11^. FET; Fisher exact test. **i**. Using logistic regression analysis, the Eigenprotein for PM13 was found to be significantly associated with incident nAMD (P-value = 1.2×10^−11^), diagnosed five years after the baseline visit. *** (P-value < 0.001).

Network-based approaches recognize that complex diseases arise from multiple proteins, genetic and environmental factors interacting in complex ways, rather than by single proteins that function alone^11,12,20-24^. The first comprehensive serum protein network, closely linked to many common diseases and under genetic control, has recently been identified^11^. This work showed that serum proteins exist in 27 coregulatory network modules, most of which are arranged in larger clusters. Many of the 28 AMD-associated proteins were correlated to each other (Supplementary Figure S5). In fact, 12 of these 28 proteins were enriched in a single module of the serum protein network (P = 2.6×10^−8^) (Figure 1h and Supplementary Table S4). This module, known as serum protein module 13 (PM13), was not associated with any of the cardio-metabolic traits previously examined^11^. However, the Eigenprotein (the first principal component) of PM13 was significantly associated with incidence of advanced AMD (nAMD, n = 150, P < 0.001) diagnosed five years after the first visit (Figure 1i), highlighting the potentially predictive nature of the network as previously stated^11^.

Serum proteins are present in distinct network modules that included proteins synthesized in all solid tissues and may serve as an integration of the body’s tissue-specific networks. This extends to AMD in that single-cell RNA sequencing analysis (Supplementary Figure S6), shows many AMD-related proteins were abundantly expressed in AMD-relevant ocular cells, whereas others were enriched in tissues such as the liver and brain (Supplementary Table S5).

### Proteins in circulation and their co-regulatory networks elucidate the genetic basis of advanced AMD

In the most recent GWAS meta-analysis of advanced AMD, Fritsche et al.^10^ examined 16,144 AMD patients, identifying 52 independent common and rare variants across 34 distinct genomic loci (Supplementary Table S6). We tested each of these variants for an effect on the 4137 proteins with the aim to narrow down the causal candidates at these genetic risk loci.

Here, 18 of the risk loci for AMD affected a total of 339 serum proteins (Supplementary Tables S6 and S7). In addition, the variant rs11080055 was linked to the previously described^11^ hotspot rs704 (*r*^2^=0.784) that affected several hundred serum proteins (Supplementary Tables S6 and S8). Intriguingly, 22 out of 28 proteins found to be associated with AMD above were controlled by one or more of six AMD susceptibility variants (Table 2).

**Table 2:**
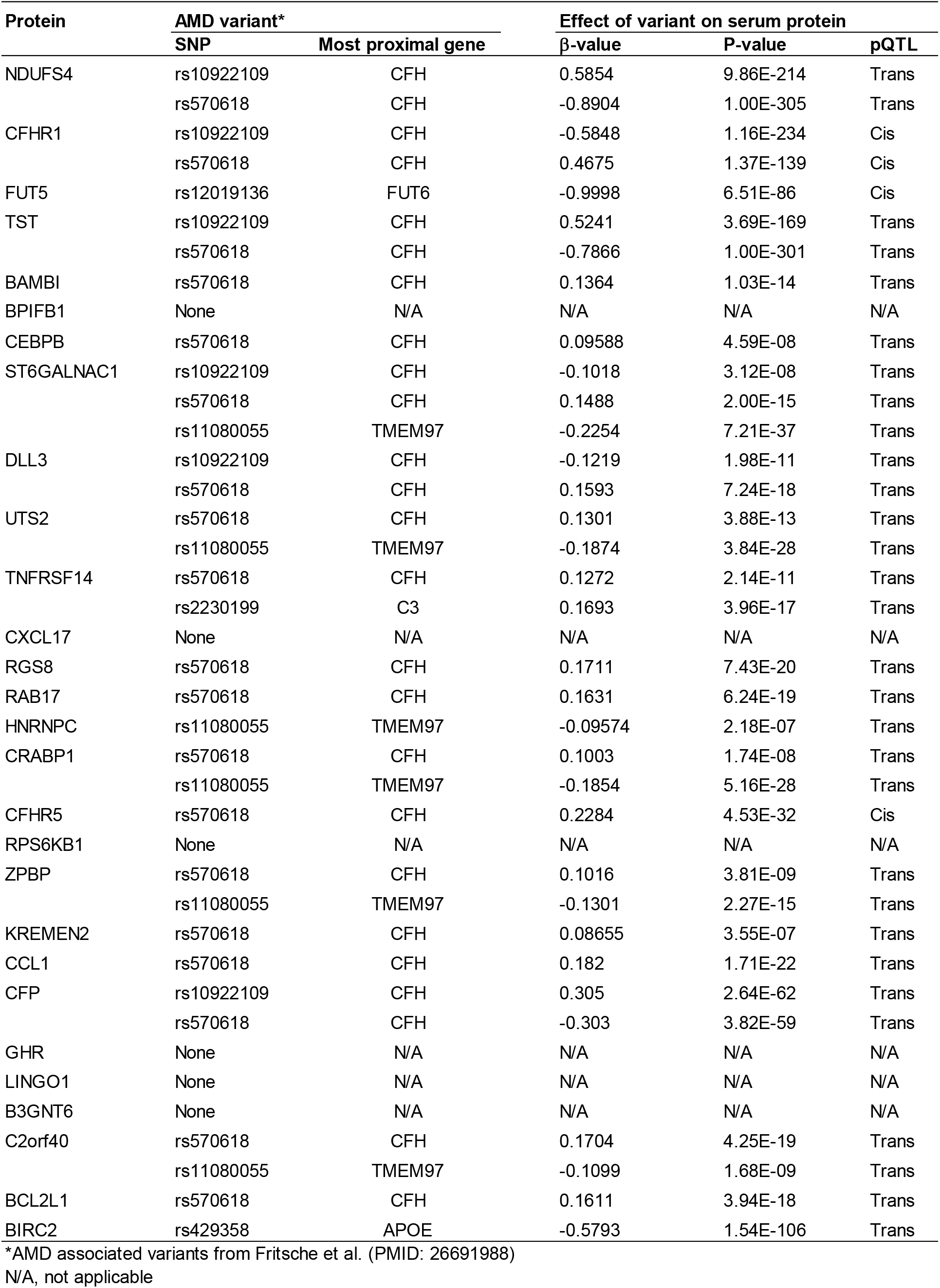
AMD-associated proteins affected by AMD risk variants.

The AMD associated variant rs10922109 in the *CFH* gene at 1q31.3, for example, was significantly associated with AMD in the AGES-RS (P = 2.6×10^−16^) (Supplementary Table S6) and influenced the levels of 40 serum proteins (Figure 2a and Supplementary Table S7), including the AMD-associated proteins CFHR1, TST, DLL3, ST6GALNAC1, CFP and NDUFS4 (Figure 2a, Table 1 and Supplementary Table S7). Among the proteins regulated by rs10922109, complement system proteins are significantly over-represented (Supplementary Table S9). These included the *cis* (proximal) regulated proteins CFHR1, CFHR4 and CFH (Figure 2b-d), as well as the *trans* (distal) regulated proteins C3, CFP (Figure 2e), and CFB. According to the GTEx database^25^, rs10922109 affects the mRNA levels of the genes encoding the *cis* regulated CFHR1, CFHR4 and CFH (Supplementary Figure S7a), with directionally consistent effects on the transcripts and cognate proteins (Supplementary Figure S7a, b). CFHR4 and CFH were not significantly associated with AMD using stringent multiple testing corrections, though the risk allele effect was directionally consistent with increased complement activation in AMD, with their relationships seen using the top and bottom quintiles and various stages of AMD (Figure 2b). Notably, CFH and the *trans* regulated protein CFP exhibit an inverse relationship with AMD outcomes compared to the CFHR1 and CFHR4 (Figure 2b). C3 and CFB, the two *trans* regulated proteins, were not linked to any AMD-related outcomes (data not shown). Finally, out of the 40 proteins regulated by rs10922109, 11 proteins cluster in protein module PM13 noted above (Supplementary Tables S10 and S11).

**Figure 2.**
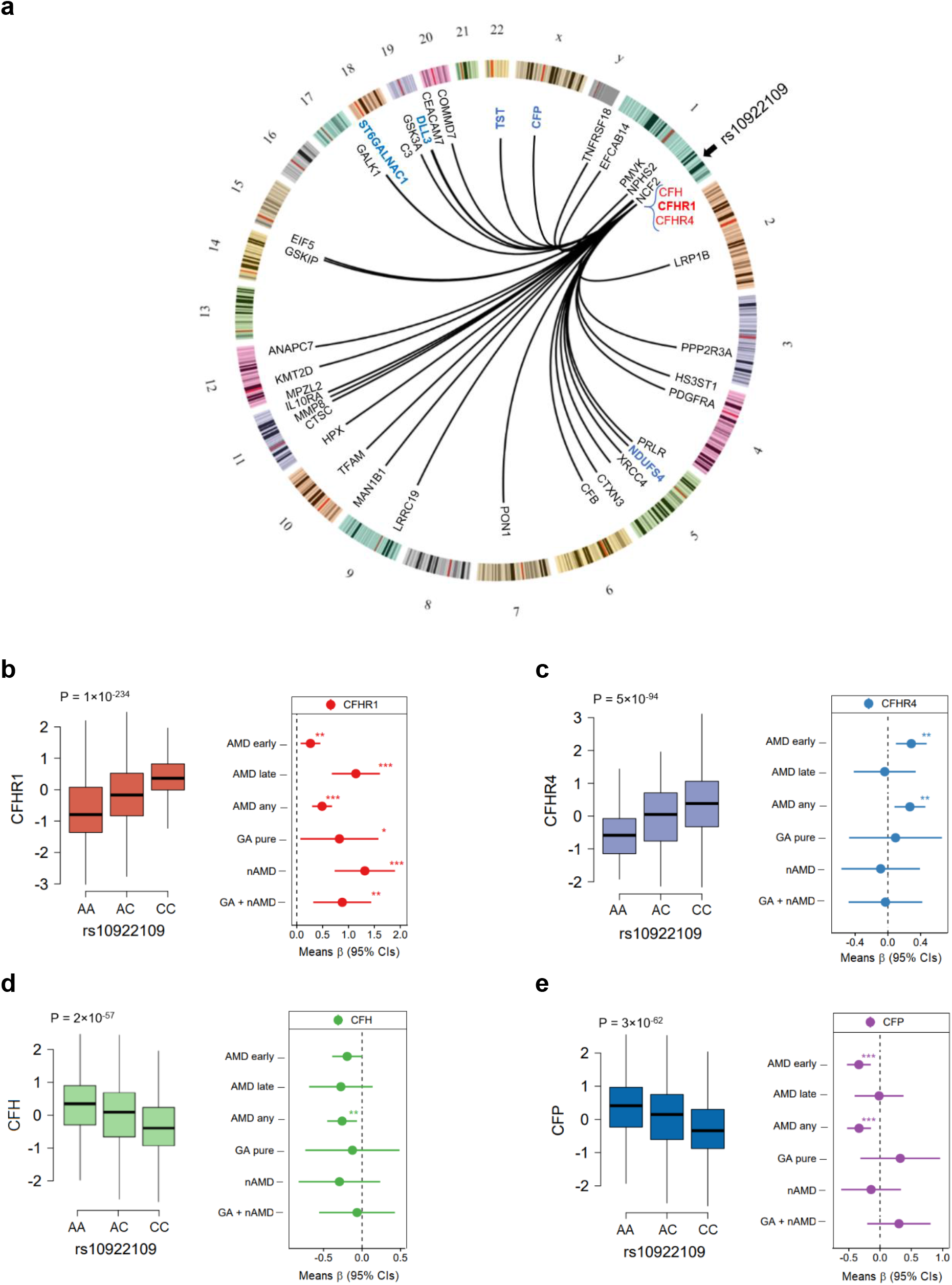
Serum proteins link genetics to AMD. **a**. The AMD-related variant rs10922109 at 1q31.3 is connected to 40 serum proteins in a Circos plot (see also Supplementary Table S7). The blue (*trans* effects) and red (*cis* effects) colored proteins are among the 28 AMD-associated proteins in Table 1. **b**. Box plot showing serum levels of CFHR1 as a function of copy C alleles for the variant rs10922109 (left panel). The relationship between the 5^th^ and 1^st^ quintile (reference) of CFHR1 serum levels and various AMD related outcomes, *** (P-value < 0.001), ** (P-value < 0.01), * (P-value < 0.05). **c**-**e**. Similar plots for CFHR4, CFH and CFP. *** (P-value < 0.001), ** (P-value < 0.01), * (P-value < 0.05).

Previously, we demonstrated that protein networks were under strong genetic control and that networks link genetics to complex diseases^11^. Consequently, we looked at the relationship between all known AMD-related GWAS variants and the 27 serum protein network modules through their Eigenproteins (1^st^ and 2^nd^ principal components). The modules in supercluster III (modules PM11 to PM15)^11^, were associated with the largest number of AMD-causing genetic variants (Supplementary Tables S12). For example, the *CFH* variant rs570618 at 1q31.3 associated with AMD risk (Supplementary Tables S6), controls 217 different serum proteins of which 100 are found in PM13 (Supplementary Tables S7 and S10). Also, rs570618 was significantly associated with the Eigenprotein for the AMD-associated protein module PM13 (Supplementary Table S12), further reinforcing its connection to AMD. In passing, rs570618 influenced 33 of the 40 proteins affected by rs10922109 (see above) and are both intronic SNPs in the *CFH* gene in a moderate linkage disequilibrium (*r*^2^ = 0.41), but the latter also regulated 184 other serum proteins (Supplementary Table S7).

### Serum proteins regulated by AMD-associated genetic variants map to core pathways involved in the pathobiology of AMD

In a recent review of the pathobiology of AMD, a comprehensive pathway map of the disease based on 110 high-risk candidate genes, was presented^26^. Many of the proteins regulated by AMD-associated variants (Supplemental Tables S7 and S8) map to these pathways, including CFH, C2, CFB, C3, CFI, PILRA and PRLR, all of which are involved in para-inflammation homeostatic processes. Others include MUC1, TIMP3, and TNXB, which are involved in extracellular matrix (ECM) homeostasis, VEGFA, involved in choroidal homeostasis, and ICAM3, in phagocytosis. VEGFA, a promoter of neo-angiogenesis, for example, is increased in the ocular tissues of nAMD patients^27^. Drusen, accumulating under the retina in AMD, are comprised of lipoprotein particles and a variety of proteins^28^, including TIMP3, CLU, VTN, in addition to complement system proteins^29-31^, many of which are regulated by the AMD-associated risk variants studied here (Supplementary Tables S6-8). For example, the *CETP* intron variant rs1864163 controlled 9 proteins, 2 of which were apoproteins (APOA5 and APOM), while rs17231506 (upstream of *CETP, r*^2^ = 0.14 with rs1864163) controlled 10 proteins, 3 of which were apoproteins (APOA5, APOM and APOC3). These findings are consistent with the response to retention hypothesis of AMD^32^. The AMD variant rs570618 in the *CFH* gene regulates CLU which is one of the most prominent proteins in drusen, and it is substantially increased in advanced AMD^33^. When the top and bottom quintiles for CLU were compared to different AMD outcomes, we found that serum CLU levels were highest in AMD with GA (Figure 3a), which is consistent with the increased levels associated with the rs570618 AMD risk allele.

**Figure 3.**
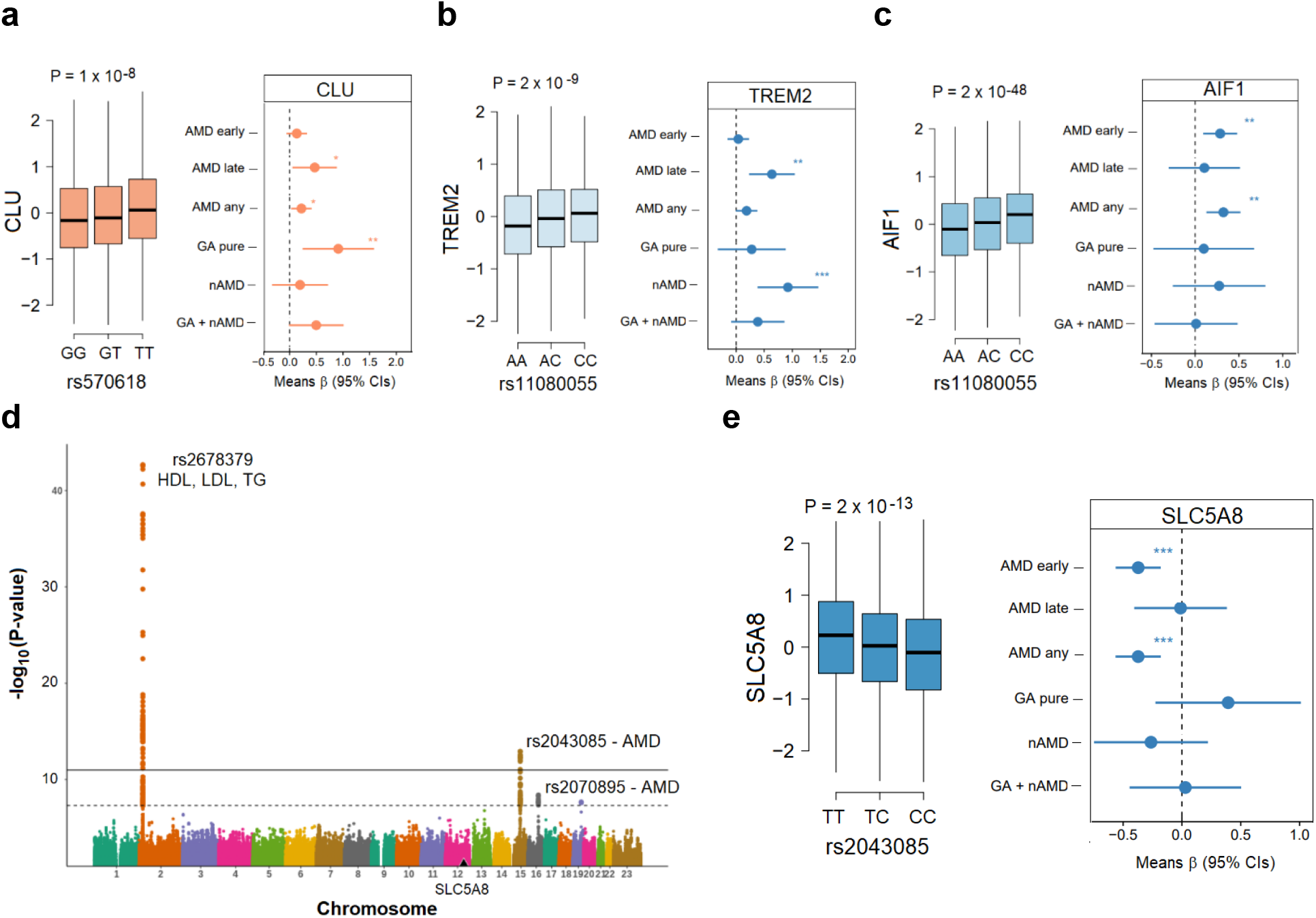
Proteins involved in various AMD-related core pathologies are linked to AMD risk variants. **a**. Box plot showing serum levels of CLU as a function of copy T alleles for the variant rs570618 (left panel). The relationship between the 5^th^ and 1^st^ quintile (reference) of CLU serum levels and various AMD related outcomes, *** (P-value < 0.001), ** (P-value < 0.01), * (P-value < 0.05). **b**-**c**. Similar plots for TREM2 and AIF1 with copy C allele for rs11080055. *** (P-value < 0.001), ** (P-value < 0.01), * (P-value < 0.05). **d**. The Manhattan plot highlights *trans*-acting variants at different chromosomes in a GWAS of serum SLC5A8 levels, specifically the AMD-associated variants rs2070895 and rs2043085, as well as the variant rs2678379, which affects lipoprotein levels. The y-axis shows the -(log_10_) of the P-values for the association of each genetic variant present along the x-axis at different chromosomes. **e**. A box plot (left panel) of the AMD-associated variant rs2043085 affecting serum levels of SLC5A8 (P = 2 × 10^−13^), and the relationship of the 5^th^ and 1^st^ (reference) quintiles of SLC5A8 levels with different AMD-related outcomes (right panel). *** (P-value < 0.001).

Our findings point to both previously implicated pathways, but also a slew of novel AMD-associated proteins and pathways. Inflammation and microglia cell recruitment has been linked to the pathophysiology of AMD, specifically retinal neovascularization^34^. The microglia associated proteins TREM2 (Figure 3b) and AIF1 (Figure 3c), were found to be controlled by the AMD risk variant and *trans* hotspot rs11080055 in the *VTN/TMEM97* locus (Supplementary Tables S8). The serum level of the microglia and macrophage activity biomarker TREM2^35^, previously linked to Alzheimer’s disease^36^, was positively correlated with nAMD and not GA (Figure 3b), implying increased macrophage (and/or microglia) activity associated with neovascularization in AMD patients’ retinas. AIF1 serum levels, like TREM2, were positively associated with an increased risk of AMD (Figure 3c). Another novel connection with AMD is SLC5A8 (aka SMCT1), a Na+-coupled transporter located in the retina^37^ that plays a role in the retina’s energy homeostasis. SLC5A8 is notable for its ability to transport 2-oxothiazolidine-4-carboxylate (OTC), a prodrug that can protect against oxidative stress, across the retina^38^. This protein was found to be regulated by two AMD-associated variants, rs2070895 and rs2043085 in or proximal to the *LIPC* gene (Supplementary Table S7 and Figure 3d), as well as the *APOB* intron variant rs2678379 (Figure 3d), which is a well-established regulator of plasma lipoprotein levels (HDL, LDL, and triglycerides)^39^. The genetic influence and disease association show that serum levels of SLC5A8 are inversely related to AMD (Figure 3e), which is consistent with the protein’s proposed protective function^37^. In summary, these findings indicate that serum proteins link genetics to the central molecular pathology of AMD in the retina.

Among the proteins affected by AMD-associated variants, categories such as the complement system, innate immunity, cell death and lipoprotein fractions, previously linked to AMD pathology^26^, were significantly enriched (Supplementary Tables S9). Proteins not previously implicated in the pathophysiology of AMD included five proteins that were regulated by rs72802342 in the *CTRB2/CTRB1* locus, enriched for pancreatic secretion (P = 1.4×10^−8^) (Supplementary Table S9). No other proteins were affected by this variant. These five proteins have been associated with pancreatitis and/or pancreatic cancer and breakdown of the ECM^40-43^. In passing, ECM pathway dysregulation is a known risk factor for AMD^10^. Finally, we find that the 339 proteins controlled by AMD risk loci (Supplementary Tables S3), were over-represented in the serum protein network PM13 and the interconnected larger cluster of modules PM11, PM14 and PM15 (Supplementary Table S12), all of which are to some extent regulated by the *CFH* locus. The inclusion of proteins regulated by the *trans* hotspot rs11080055, located at the *VTN/TMEM97* locus, in the immune and inflammation related cluster of modules has been previously reported^11^. It should be noted though, that rs11080055 only accounts for a very small portion of AMD’s genetic liability (Supplementary Table S6).

### Identifying causal candidate proteins for AMD using two-sample Mendelian randomization analysis

To determine if any of the 28 AMD-associated proteins might be causally related to the disease, a two-sample Mendelian randomization (MR) study was performed using *cis*-acting genetic variants as instruments. BPIFB1, CFHR1, CFHR5, FUT5, and GHR were found to have such instruments. The causal estimate for each protein was determined using the generalized least squares method (GWLS)^44^, where CFHR1, CFHR5 and FUT5 were found to be significant (FDR < 0.05) (Figure 4). For both definitions of AMD (see Methods), the proteins CFHR1, CFHR5, and FUT5 were found to be significant and directionally consistent with their corresponding observational estimates (Figure 4), highlighting their role as risk factors for AMD development. Of the three proteins, CFHR1 was detected with ELISA in a much smaller sample of an independent study of healthy volunteers and AMD patients, confirming elevated circulating levels of CFHR1 in the disease’s most advanced stage (Supplementary Figure S8 and Methods). In a secondary analysis, we repeated the MR analysis, but this time we included all 1327 aptamers with *cis*-acting genetic instruments. In this study, 21 additional proteins, such as ADAM19, C3, CFI, AIF1, and VTN, were found to have a significant causal estimate for AMD (Supplementary Figure S9). These proteins were collectively enriched for the complement and coagulation cascade (FDR = 0.00002). Different aptamers for C3 and VTN produced opposite effects, potentially due to aptamers binding different protein epitopes or isoforms (Supplementary Figure S9). Both CFHR1 and FUT5 remained statistically significant at this more stringent multiple testing correction threshold (Supplementary Figure S9).

**Figure 4.**
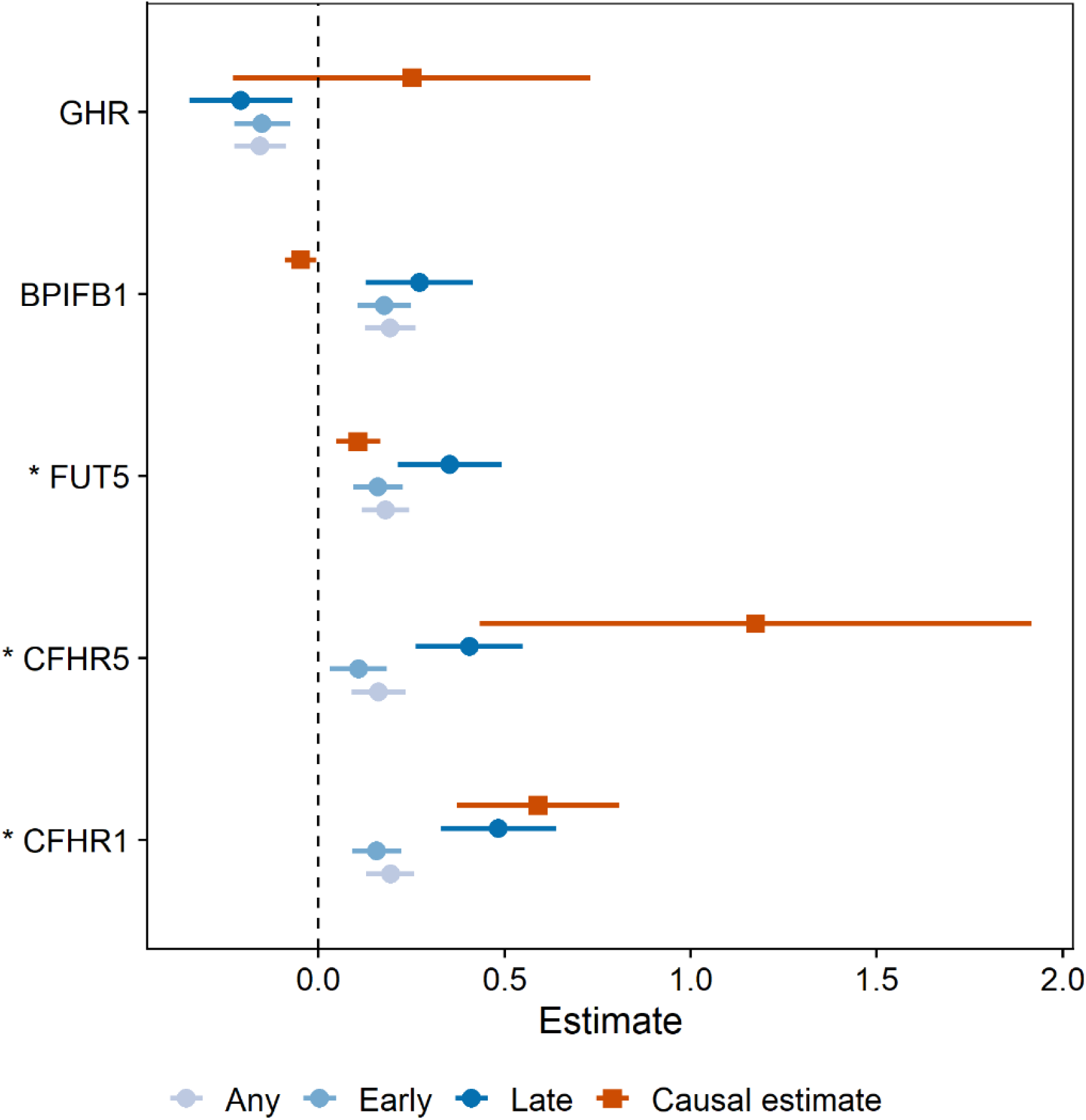
A two-sample MR analysis of the 28 AMD-associated proteins. The causal estimate (red squares) from the two-sample MR analysis compared to the observational estimates (circles) for each of the five proteins with *cis*-acting instruments and associated with AMD in the observational study. As each protein could have different observational coefficients depending on which definition of AMD (see Methods) was used, it was decided to select and display the coefficient for each definition which had the lower adjusted P-value. The direction of the causal estimate and observational estimates were consistent for proteins CFHR1, CFHR5 and FUT5 and inconsistent for proteins GHR and BPIFB1. The causal estimator for CFHR1, CFHR5 and FUT5 was significant (FDR<0.05) and positive, indicating that an increase in the serum level of these proteins increase the risk of developing AMD. * (FDR < 0.05).

## Discussion

Homeostasis, the maintenance of a global state, has long been recognized as a property of living systems and is maintained by integrating local and global signals so that each tissue does not act independently, but rather influences and responds to dynamically preserve the biologic equilibrium. The measurement of the levels of thousands of proteins in serum of thousands of individuals has begun to illuminate the process by which homeostasis may occur^11,12,17^. In this study we document how serum proteins report on and influence AMD appearance and progression in the eye. Previous studies of AMD patient plasma and urine reveal changes in lipid and energy metabolites however sample size has limited detection of serological protein changes, or stratification of late AMD into GA and nAMD^45-50^. The population-based AGES study with its array of biomarkers, clinical profiles, and genetic risk factors collected prospectively from participants who were aged >67 at baseline visit, and a follow up visit after five-years has enabled us to identify circulating proteins and protein networks in patients that associate with AMD stage and progression.

Consistent with the known links between AMD and the complement pathway, three complement pathway proteins were associated with AMD (CFHR1, CFHR5 and CFP). Six additional proteins known to modulate both the innate and adaptive immune response were also associated with AMD (TNFRSF14, CCL1, CXCL17, BPIFB1, BIRC2 and CEBPB), implicating broad induction of inflammatory processes in AMD. Reductions in two mitochondrial proteins, the complex 1 protein NDUFS4 and the sulfotransferase protein TST were also associated with AMD, consistent with structural, functional, and genetic mitochondrial changes in AMD^51-55^.

Inactivation of the NDUFS4 gene causes a severe form of the blinding disease Leigh syndrome, and mice lacking this gene die prematurely at 50-60 days old^56^, with compromised photoreceptor function^57^ and excessive lipid droplet formation^58^. Elevation of the ribosomal S6 protein kinase 1 (RPS6KB1), a component of the nutrient-responsive mTOR (mammalian target of rapamycin) signaling pathway was also associated with AMD, consistent with the AMD-like phenotype in mice with RPE mTOR overactivation^59-61^. The pro-longevity effects of *RPS6KB1* gene deletion^62^, or mTORC1 inhibition across multiple species^63^, are also intriguing when considering AMD as a disorder of aging.

The rate of progression from early to late AMD was ∼4.5% per year, with 12.7% progressing to GA and 7.6% to nAMD after 5 years. Serum levels of PRMT3 protein were elevated in early AMD patients progressing to GA, but not in those progressing to nAMD. PRMT3 controls ribosomal activity *via* arginine methylation of the 40S ribosomal subunit protein RPS2^64-66^.

Interestingly, control of mRNA translation is *via* phosphorylation of another component of the 40S ribosomal subunit, RPS6, by the previously mentioned kinase RPS6KB1, whereas another ribosomal 40S subunit RPS10 was controlled by the AMD-associated variants rs570618 and rs429358. Alterations in post-translational regulation of protein synthesis by both phosphorylation and methylation could be a key early driver of AMD pathogenesis. We also examined the association of protein networks with AMD, since deep profiling has revealed the modular structure of the serum proteome containing 27 distinct protein modules^11^. Interestingly, the protein module PM13 is linked to incident nAMD and is enriched with individual proteins linked to AMD (12 of 28) and/or proteins regulated by AMD risk variants. Serum protein network changes are highly predictive of overall survival and disease progression in complex diseases such as cardiovascular and metabolic diseases^11^, and these data demonstrate the ability of serum protein modules to also predict advanced AMD.

AMD is associated with 34 distinct genomic loci, accounting for nearly half of AMD’s genetic liability^10^. Integrating intermediate traits such as mRNA and/or protein levels with genetics and disease traits helps in identifying the causal candidates^11,21-24^. The present study integrated the most recent findings of AMD genetic risk factors with 4137 human serum proteins, providing mechanistic insights into previously described AMD SNPs. Many of the variants that explained the largest fraction of the genetic liability for AMD, regulated a wide range of circulating proteins. Interestingly, of the AMD-associated proteins identified, the majority were controlled by one or more of only six AMD susceptibility variants. For example, the *CFH* variant rs10922109 (C), a strong risk factor for AMD (OR = 2.63), increases levels of the CFHR1 and CFHR4 proteins, while also decreasing CFH. Consistent with these changes in protein levels, rs10922109 (C) has previously been associated with *activation* of the complement cascade in AMD patients^67^ and also increased serum CFHR4 by orthogonal methods^68^. AMD variants had both overlapping and distinct effects on serum proteins and controlled numerous proteins in the complement cascade. For instance, the *CFH* variant rs570618 influenced 33 of the 40 proteins affected by the co-localized variant rs10922109 (e.g., CFHR1/4, C3, CFB and CFP), but rs570618 also regulated 184 other serum proteins (e.g., CFHR5, C1S and C8) (Supplementary Table S7). It should be noted that the AMD risk allele for the rs570618 variant was not linked to higher levels of complement activation in AMD patients^67^ implicating non-complement pathways associated with this variant (Supplementary Table S9), or increased complement activity at ocular cell surfaces such as Bruch’s membrane or choriocapillaries^67^.

Beyond complement, variant rs62358361 (G), located in an *C9* intron, which is protective for AMD, significantly reduces CREBBP – the master regulator of hepatocyte lipid metabolism, implicating a potential role for liver metabolism pathways in AMD. In contrast to these strong genetic associations driving systemic protein changes, rs3750846 (OR 2.81) in the *ARMS2/HTRA1* locus on chromosome 10 does not affect serum protein levels in the present study. One reason could be that the current platform lacks aptamers that detect the ARMS2 and HTRA1 proteins encoded by genes near rs3750846, but the variant is known to regulate the transcription of both *ARMS2* and *HTRA1* in solid tissues^69^. Alternatively, this variant may alter ocular specific protein changes that are not apparent in serum.

CFHR1 and CFHR5 are presumed pathogenic since they impede CFH binding to pro-inflammatory lipid peroxidation products^70,71^, and induce inflammasome activation^72^, whereas CFHR1 gene deletion is known to be protective for AMD^70^. Indeed, the two-sample MR test analysis revealed that both proteins could be causally linked to AMD. However, because their *cis* regions overlap (Supplementary Table S7), from which the genetic instruments are selected, it is impossible to say whether one or both are the causal effector. In addition, the FUT5 was supported as causal candidate in the MR analysis, which was also strongly associated with risk of AMD (Table 1). As 23 of the AMD-associated proteins did not have any *cis*-acting instruments, they could not be tested in the MR analyses, and we thus cannot exclude their candidacy as causal proteins. In the extended MR analysis, 21 additional proteins were supported as causal, including multiple other members of the complement system (CFI, C3, C1R, F11, VTN), further highlighting the causal role of this pathway in the development of AMD.

AMD is a multifactorial age-related disease with a complex pathobiology in which systemic and local inflammatory and other effectors play significant roles. Recent evidence indicates that thousands of proteins present in serum participate in cross tissue regulation that connects all parts of the body^11,12^. This occurs by tissues releasing protein(s) into the bloodstream which control biological processes in other physically distant tissues, resulting in a network of cross-tissue regulatory loops^12^. For example, the expression of the proteins CFHR1 and CFHR5, which have been causally linked to AMD in this study (Figure 4), are highly specific to the liver, whereas FUT5 is expressed in the bone marrow and testis (Supplementary Table S5). More research is required to determine the roles of the numerous serum proteins discussed in this study and their potential effect on various pathophysiological processes that lead to various stages of AMD in the local eye setting. Many of the serum proteins associated with AMD in this study are characterized as intracellular proteins, and the significance of their presence in serum, remains to be determined. Our findings provide a robust framework for understanding the pathobiology of AMD, which may lead to the discovery of new systemic biomarkers and therapeutic targets.

## Methods and Material (*online content*)

### Study population

Participants aged 66 through 96 were from the Age, Gene/Environment Susceptibility Reykjavik Study (AGES-RS) cohort^73^. AGES-RS is a single-center prospective population-based study of deeply phenotyped subjects (5764, mean age 76.6±5.6 years) and survivors of the 40-year-long prospective Reykjavik study (n~18,000), an epidemiologic study aimed to understand aging in the context of gene/environment interaction by focusing on four biologic systems: vascular, neurocognitive (including sensory), musculoskeletal, and body composition/metabolism. Descriptive statistics of this cohort as well as detailed definition of the various disease endpoints and relevant phenotypes measured have been published^11,73^. Of the 5764 AGES-Reykjavik participants 3411 attended a 5-year follow-up visit (AGES-RS II). The AGES-RS was approved by the NBC in Iceland (approval number VSN-00-063), and by the National Institute on Aging Intramural Institutional Review Board, and the Data Protection Authority in Iceland.

Detailed description of AMD diagnosis and the baseline characteristics of the AMD population in AGES-RS has previously been described in two separate publications^74,75^, which vary in terms of the medical definition of early stage AMD. In Holliday et al.^75^, early stage AMD (n = 1755) was defined as the presence of soft drusen (>63 µm) alone, retinal pigment epithelium (RPE) depigmentation alone or a combination of soft drusen with increased retinal pigment and/or depigmentation in the absence of late AMD, while Jonasson et al.^74^ defined early AMD (n = 1054) by the presence of any soft drusen and pigmentary abnormalities (increased or decreased retinal pigment) or the presence of large soft drusen ≥125µm in diameter with a large drusen area >500µm in diameter, or large ≥125µm indistinct soft drusen in the absence of signs of late AMD^74^. Late AMD (n = 272) was defined by the presence of any of the following: geographic atrophy (GA) or exudative AMD including subretinal hemorrhage, subretinal fibrous scar, RPE detachment, or serous detachment of the sensory retina or signs of treatment for neovascular AMD^74^. Also, early and late AMD was combined as AMD any. Late AMD was separated into GA pure (n = 112) or neovascular AMD (nAMD) pure (n = 160), but was also combined for any form of GA as GA + nAMD (n = 183). We used both medical definitions of early AMD (and AMD any) separately to analyze the association of serum proteins with AMD.

### Protein measurements via SOMAmers

For the AGES-RS, we used a distinct version of the SomaScan platform (Novartis V3-5K), based on the slow-off rate modified aptamer (SOMAmer) protein profiling technology^11,12,76^. The aptamers are small single-stranded 40-mer DNA oligomers with modified nucleic acids selected to specifically recognize target proteins in their native three-dimensional state and show slow dissociation kinetics (t_1/2_>30 min) which in combination with stringent wash steps impedes nonspecific binding^76^. The custom-design SOMAscan platform was built to quantify 5034 protein analytes in a single serum sample with a focus on proteins that are known or expected to be present extracellularly or on the surface of cells, 4782 of which SOMAmers directly bind to 4137 different human proteins. Blood samples were collected at the AGES-RS baseline, after an overnight fast. Serum was prepared using a standardized protocol^77^, stored in 0.5 ml aliquots at -80°C and serum samples that had not been previously thawed were used for the protein measurements. To avoid batch or time of processing biases, the order of sample collection and processing for protein measurements were randomized and all samples run as a single set at SomaLogic Inc. (Boulder, US). All SOMAmers that passed quality control had median intra-assay and inter-assay coefficient of variation (CV) < 5% or equivalent to reported variability^78^. Various metrics, including both aptamer specificity direct tandem mass spectrometry (MS) analysis and inferential assistance using genetic analysis, have been used for the performance of the proteomic platform, suggesting strong target specificity throughout the platform^11^. Hybridization controls were used to correct for systematic variability in detection and calibrator samples of three dilution sets (40%, 1% and 0.005%) were included so that the degree of fluorescence was a quantitative reflection of protein concentration. Box-Cox transformation was applied on the protein data^79^, and extreme outlier values excluded, defined as values above the 99.5th percentile of the distribution of 99th percentile cutoffs across all proteins after scaling, resulting in the removal of an average 11 samples per SOMAmer. Previous studies have shown that pQTLs replicate well across different study populations as well as proteomic platforms^11,19^. While a recent comparison of protein measurements across different platforms showed a wide range of correlations^80^, *cis* pQTLs detection and validation by orthogonal MS-based measures were predictive of a strong correlation across platforms and were great indicators of platform specificity when protein concentrations obtained by orthogonal methods differ. The aptamer specificity of six of the AMD-associated proteins listed in the main text has already been validated by orthogonal mass spectrometry (MS)-based approach and/or inferred measures by proximal *cis* variants assessment for SOMAmer-protein interactions^11^. Nine additional aptamers were confirmed with SOMAmer pull down and MS (SP-MS) using AMD patient serum samples (Supplementary Table S13). There are many reasons why some aptamer-enriched proteins are not detected by SP-MS. For instance, MS analyses have different sensitivity than that of SOMAmer scan and is dependent on the protein sequence. Factors such as ion suppression of individual peptides caused by instrument/interfering molecules, incomplete trypsin digestion, and modifications of peptides either naturally or artifactually can limit the detection.

### Protein measurements via ELISA

Participants aged 55 and over (up to n=60, 15 per group), enrolled in a prospective study by Ophthalmic Consultants of Boston, to measure complement and disease-related protein biomarkers in blood. AMD patients with Age-Related Eye Disease Study (AREDS)^81^ grade 2-4 had best corrected visual acuity (BCVA) of at least 20/200. Healthy volunteers were aged matched (+/-2 to AMD patients) with BCVA of 20/40 or better in both eyes, and a comprehensive eye examination within previous 12 months revealing no diagnosis other than refractive error, mild cataract, dry eye or AREDS grade 1. AREDS grading criteria for grade 2 included mild changes including multiple small drusen, non-extensive intermediate drusen, and/or pigment abnormalities. AREDS grade 3 included at least 1 large drusen of at least 125 µm in diameter, extensive intermediate drusen, and/or noncentral geographic atrophy.

AREDS grade 4 included advanced AMD central geographic atrophy in one or both eyes. Qualifying subjects completed an informed consent form, and no identifying patient information was included. A blood sample of 10 milliliters was collected by routine phlebotomy, in EDTA-coated, lavender-topped collection tubes. Plasma was isolated after centrifugation at 4,000 rpm for 30 minutes, frozen and stored at -80°C. Rabbits were immunized with recombinant human FHR1 protein at Covance (Princeton, NJ), and CFHR1 cross-reactive antibodies were purified from serum using CFHR1-conjugated CNBr-activated Sepharose 4B resin (Cytiva, Marlborough, MA). Additionally, antibodies showing cross-reactivity to Factor H were removed by several rounds of depletion using Factor H-conjugated resin. CFHR1 levels in patient plasma were measured in a Meso Scale Discovery assay (Rockville, MD) using the selective antibodies.

### Analysis of gene expression in single cell RNA sequencing data from eye tissues

Normalized single cell data was downloaded from GEO (GSE135922) and was analyzed with the R package Seurat (v.3.0.0) in R 3.6.3 environment. Final dataset contained 4335 cells after filtering.

Variable genes were identified using Seurat with default parameters and Principal Components Analysis (PCA) was performed on these variable genes. First 11 PCs of the single cell data (resolution =0.2) were used for clustering cells with similar gene expression profile. Clusters were identified using FindNeighbors and FindClusters functions from Seurat package and UMAP dimensionality reduction was utilized for cluster visualization. The cell clusters were then manually annotated based on the markers reported in the paper^82^.

### Statistical and genetic analysis

We used linear or logistic regression for the associations of individual proteins as well as Eigenvectors of protein modules with various phenotypic measures, depending on the result being continuous or binary. Statistical results were obtained using linear models for continuous outcomes and generalized logistic models for binary outcomes for comparison of protein quintiles and AMD-related clinical traits. The models were fit using the outcome phenotypes as dependent variables and protein quintiles as predictor variables along with the adjustment variables age and sex. The protein quintiles were treated as factor variables to avoid underlying assumptions regarding linearity of effects. Continuous outcomes were standardized using z-scores prior to model fitting thus coefficient estimates should be interpreted on the standard deviation scale. That is an estimated mean difference of 1 between protein quintiles translates to a one-standard-deviation difference between groups after adjusting for other included variables. The expected means were obtained as linear predictions from the fitted models along with the fitted confidence intervals around the mean. The linear predictions for qualitative phenotypes are shown on the log-odds scale. The difference between the 5th and 1st protein quintiles was obtained as the expected marginal difference between those groups. Thus, for continuous outcomes, they are the optimal linear estimator with corresponding confidence intervals and P-values, but for discrete outcomes they are obtained using the commonly applied asymptotic approximations.

We applied linear regression using an additive genetic model for all single-point SNP association analyses for different disease-related outcomes. The results of associating genetic variants with serum protein levels were obtained using a GWAS of 4137 human proteins measured in serum using 7,506,463 assayed and imputed genetic variants from 5457 AGES-RS individuals^83^. All statistical analyses of the present study were conducted using the software environment R statistical package version 3.6.0 (2019-04-26).

### Two-sample Mendelian randomization analysis

In a two-sample Mendelian randomization study, genetic variants for each trait, the protein-encoding gene (exposure) *X* and AMD (outcome) *Y* are found in two distinct samples. A genetic variant (SNP) *Z* is used in the analysis provided it fulfills the three assumptions of instrumental variables:

1. There exists a significant association between SNP *Z* and exposure *X*.
2. SNP *Z* is independent of any confounder *U* which might influence exposure *X* and outcome *Y*.
3. SNP *Z* is independent of outcome *<u>Y</u>* conditional on exposure *X* and confounder *U*. This assumption is usually referred to as the *exclusion criterion*.

The first assumption is readily tested by setting a threshold on the significance level on the SNP-exposure association. Unfortunately, it remains a challenge to examine the validity of the second and third assumptions. When a SNP violates the third assumption, we generally speak of *pleiotropy*. To obtain the SNP-exposure associations, all genetic instruments within a 1 Mb (± 500kb) *cis* window for the protein-encoding gene were obtained for a given SOMAmer. A cis-window-wide significance level P_b_ = 0.05/N, where N was the number of SNPs within a given cis-window, was computed. Genetic instruments within the *cis* window for each SOMAmer were then clumped such that variants in high linkage disequilibrium (LD) (r^2^ ≥ 0.2) within a 1 Mb region were combined. The list of variants was then further pruned by removing all instruments with P ≥ P_b_.

Summary statistics on the genetic risk on AMD were obtained from a GWAS provided by the IAMDGC consortium^10^. Any SNP in the *cis* window-wide significant data set not found in the AMD GWAS data set were replaced by proxy SNPs (r^2^ > 0.8) when possible, to maximize SNP coverage. Causal estimate for each protein was obtained by the generalized weighted least squares (GWLS) method^44^, which accounts for the correlation that can exist between instruments. Let *Z* = {1,2, …,*M*}be an index set of all SNPs associated with a protein-encoding gene and AMD. For *j* ϵ *Z*, denote the *j*-th SNP-protein association and SNP-AMD association as 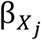 and 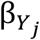 respectively, each with their corresponding standard error 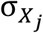 and 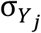. Then, the causal estimate 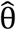 is found by evaluating:

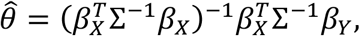

where 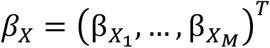 and 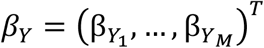 and Σ^-1^ is the inverse of the weighting matrix Σ whose *(i, j)*-th entry is 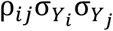 and ρ_*ij*_ is the correlation between SNPs *i, j* ϵ *Z*. The standard error of 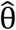 is 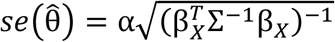, where 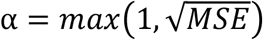 with:

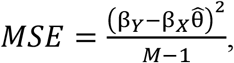

reflects our uncertainty about the weights by assuming that we only know their relative magnitudes.

Finally, each estimate was subjected to a two-step sensitivity analysis. Any protein with a causal estimate found to be significant after adjusting the P-value with the Benjamini-Hochberg method was reassessed with the weighted median estimator which allows for up to 50% of the genetic instruments to violate any of the three assumptions of instrumental variables^84^. If the direction of the weighted median estimator was consistent with the GWLS estimate and remained significant at P < 0.05 it was subjected to the second stage of the sensitivity analysis but otherwise removed. Any protein which passed the first step was then reexamined with MR-Egger estimator which replaces the exclusion criterion with a weaker condition, the InSIDE assumption^85^, which allows SNPs to exhibit pleiotropic effects provided that the pleiotropic effect is uncorrelated with the SNP-AMD effect 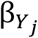. As with the first step, any protein whose MR-Egger estimate was directionally consistent with the GWLS estimate and significant at P < 0.05 was kept and considered a causal candidate. Causality for proteins with single *cis*-acting variants was assessed with the Wald ratio estimator 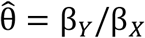.

## Data Availability

The custom-design Novartis SOMAscan is available through a collaboration agreement with
the Novartis Institutes for Biomedical Research. Data from the
AGES Reykjavik study are available through collaboration
under a data usage agreement with the IHA. All data supporting the conclusions of the paper are presented in the main text and supplementary materials.

## Acknowledgements

We thank the IAMDGC consortium for supplying us with their GWAS summary statistics data. The Age, Gene/Environment Susceptibility-Reykjavik Study (AGES-RS) was supported by NIH contracts N01-AG-1-2100 and HHSN27120120022C, the NIA Intramural Research Program, Hjartavernd (the Icelandic Heart Association), and the Althingi (the Icelandic Parliament). V.E. and Va.G. are supported by the Icelandic Research Fund (IRF grants 195761-051, 184845-053 and 206692-051).

