## Supplementary Figures S1-S9 for "A Proteogenomic Signature of Age-related Macular Degeneration in Blood"

[1]Icelandic Heart Association, Holtasmari 1, IS-201 Kopavogur, Iceland.
[2]Faculty of Medicine, University of Iceland, 101 Reykjavik, Iceland.
[3]Novartis Institutes for Biomedical Research, 22 Windsor Street, Cambridge, MA 02139, USA.
[4]GNF Novartis, 10675 John Jay Hopkins Drive, San Diego, CA 92121, USA.
[5]Department of Ophthalmology, University Hospital, Reykjavik, Iceland.
[6]Laboratory of Epidemiology and Population Sciences, National Institute on Aging, MD, USA.
[7]Division of Epidemiology and Clinical Applications, National Eye Institute, National Institutes of Health, Maryland, USA.

#### Figure Legends

**Supplementary Figure S1**. **Association of global serum proteins with different stages of AMD outcome**. **a.** Using logistic regression analysis and Bonferroni correction for multiple comparisons, a volcano plot of all serum proteins associated with AMD early is shown, with colored data points highlighting significant associations. Definition of early AMD was according to Jonasson et al.[1]. **b**. A volcano plot of late AMD, and **c**. A volcano plot of AMD any, where early stage of the disease wase defined according to Jonasson et al.[1].

**Supplementary Figure S2**. **Protein levels changing from no-AMD to advanced AMD**. **a**. A violin plot showing a near steady rise in levels of CFHR1, **b**. CFHR5 and **c**. BPIFB1 from no AMD to early AMD to advanced nAMD.

**Supplementary Figure S3**. **Quintiles of all 28 AMD-associated protein levels in relation to various AMD-related outcomes**. The relationship between the different quintiles for the 28 AMD-associated protein levels and the various AMD-related outcomes (see Methods for details). The x-axis represents the quintiles for each protein, and the y-axis represents the estimate (Methods).

**Supplementary Figure S4**. **Estimated difference between the top versus bottom quintiles of the AMD-associated proteins in relation to various AMD outcomes.** Association of the top and bottom quintiles of AMD-linked protein levels to the various AMD related outcomes (see Methods for details). ***(P-value < 0.001), **(P-value < 0.01), *(P-value < 0.05).

**Supplementary Figure S5. Correlation between the 28 AMD-associated proteins.** A heatmap showing the Spearman rank inter-correlations of all 28 AMD-associated proteins, revealing a large cluster of inter-correlated proteins.

**Supplementary Figure S6**. **Single cell RNA sequencing analysis of the eye.** RNA sequencing was used to examine the expression of the various AMD-associated proteins described in the current study in the RPE choroid (macula), at a single cell level, from publicly available dataset (GSE135922)[2].

**Supplementary Figure S7**. **mRNA and protein changes due to rs10922109 variant**. **a**. Violin plot showing mRNA levels of CFHR1, CFHR4 and CFH as a function of copy C alleles for the variant rs10922109. The data came from the Genotype-Tissue Expression (GTEx) portal. **b**. Box plot showing serum levels of CFHR1, CFHR4 and CFH as a function of copy C alleles for the variant rs10922109.

**Supplementary Figure S8**. **Protein levels changing from no-AMD to advanced AMD**. **a**. A violin plot showing increasing nM concentrations of CFHR1 from no AMD to early AMD to advanced nAMD. Here, AMD patients had AREDS grade 2-4, while AREDS grade 1 are healthy aged-matched volunteers (Methods). Protein concentrations were determined by ELISA. ***(P-value < 0.001), **(P-value < 0.01).

**Supplementary Figure S9**. **Proteins identified as significant causal candidates for AMD in the MR analysis.** Significant causal estimates and their 95% confidence intervals for proteins with a) multiple genetic instruments (GWLS) and b) proteins with a single genetic instrument (Wald ratio) after adjusting the P-value for all 1327 aptamers with *cis*-acting variants with the Benjamini-Hochberg method. CFHR1 and FUT5 were identified as potential risk factors for developing AMD in both the MR and observational analysis. FCN3, C3 and VTN appear twice as each had two significant aptamers. C3 and VTN produce opposite effects for each of their aptamers.

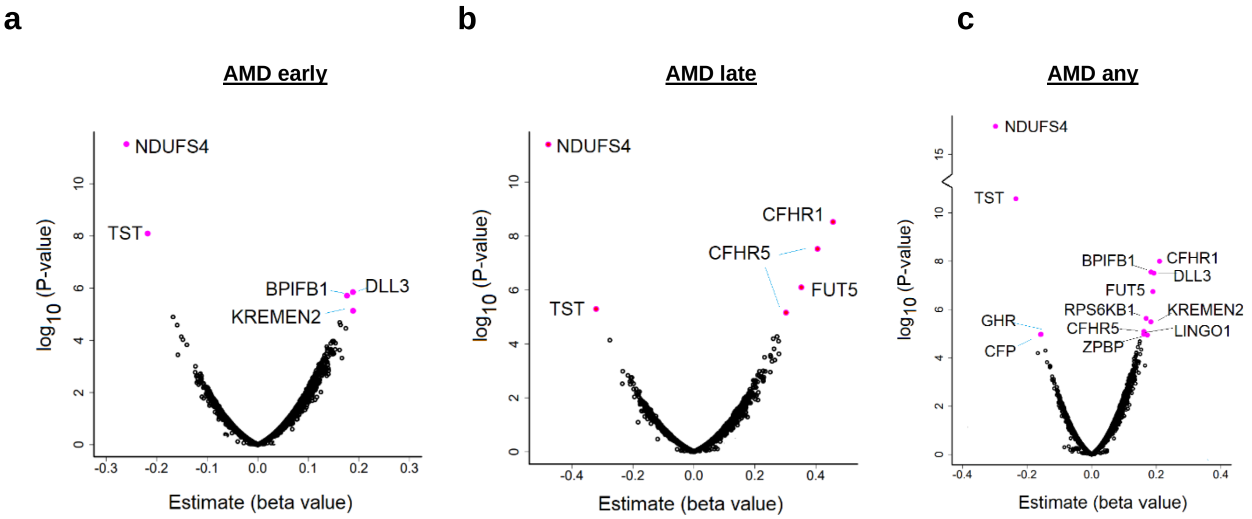


### Supplementary Figure S1

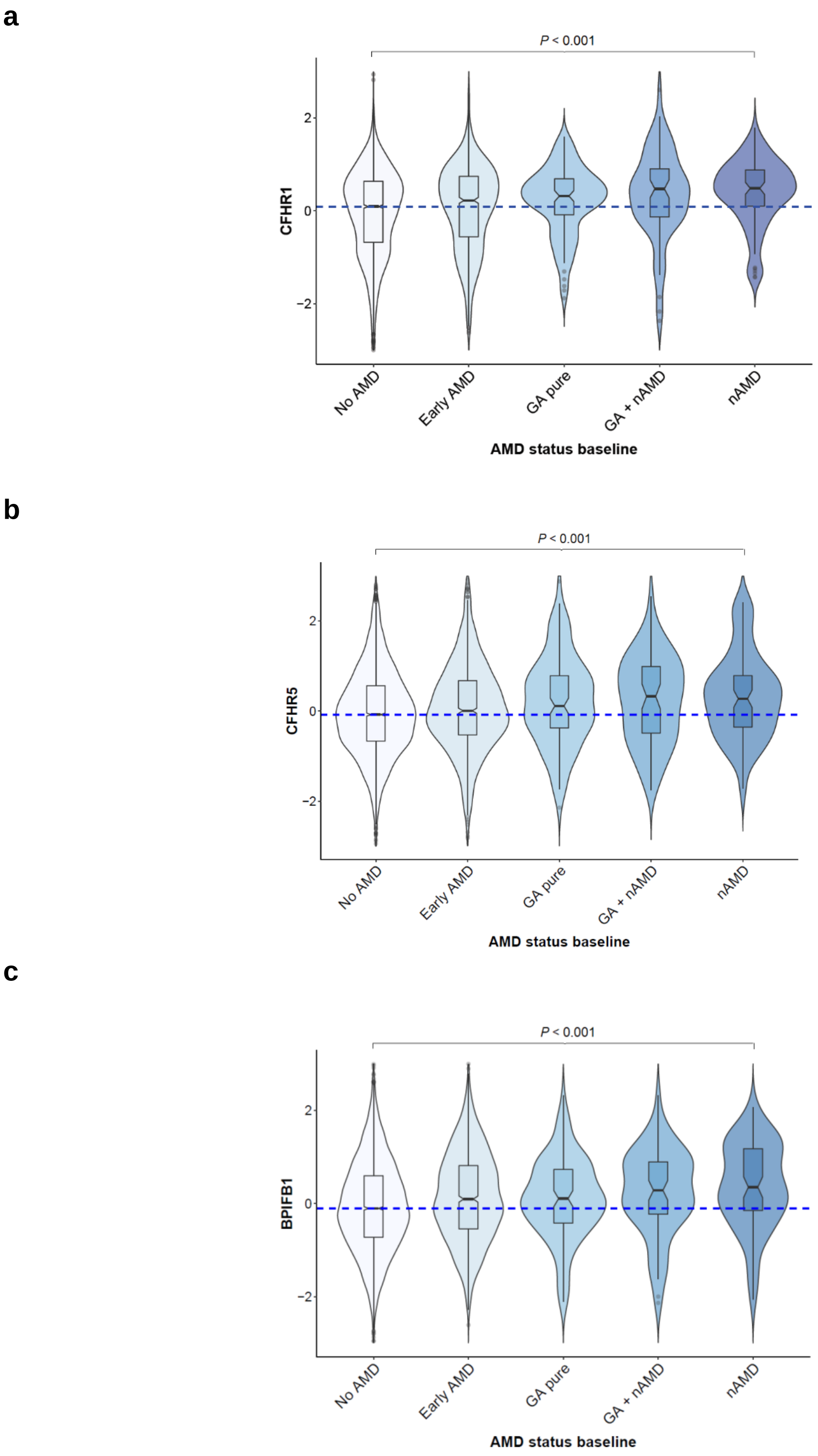


### Supplementary Figure S2

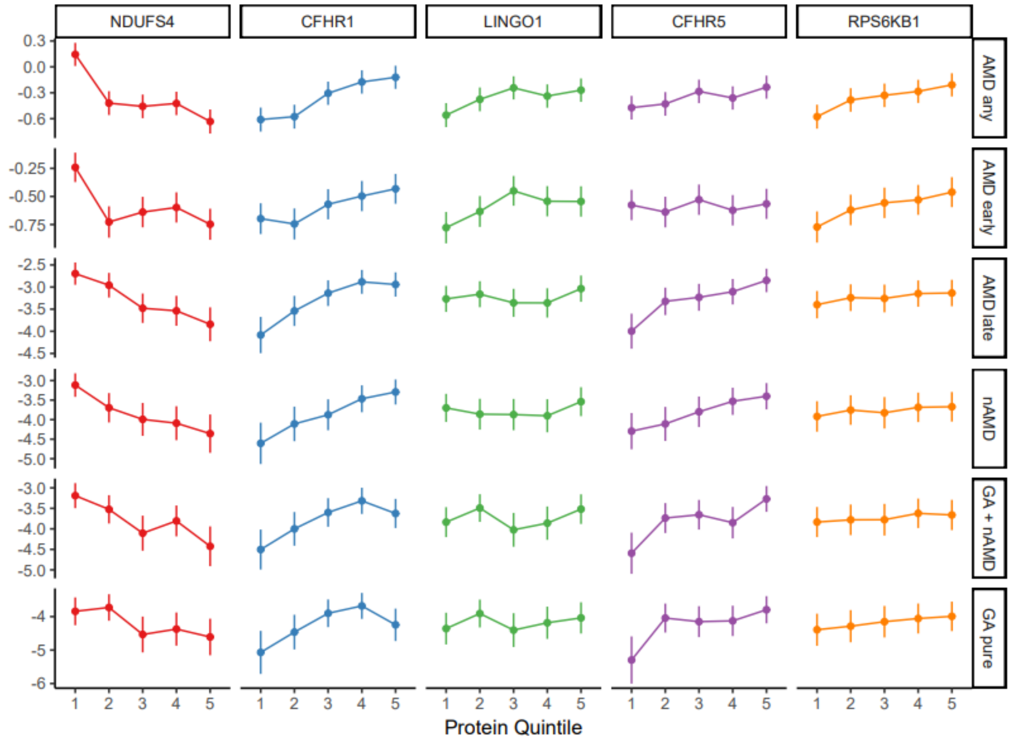
NDUFS4
CFHR1
LINGO1
CFHR5
RPS6KB1
AMD any
AMD early
AMD late
nAMD
GA + nAMD
GA pure
Protein Quintile

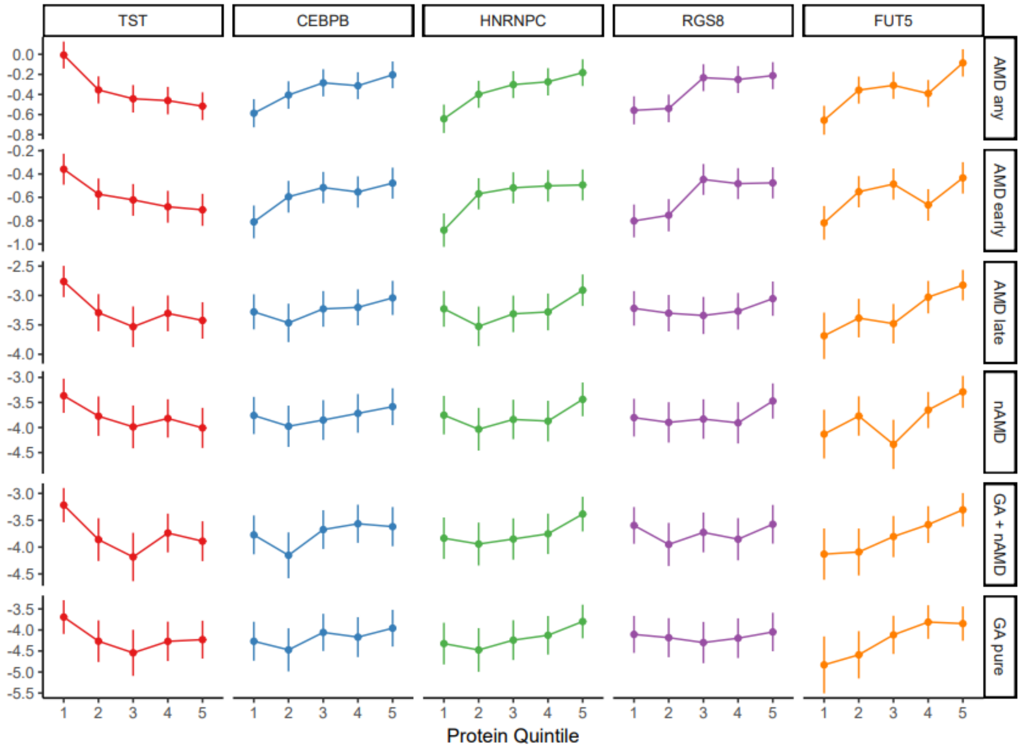
TST
CEBPB
HNRNPC
RGS8
FUT5
AMD any
AMD early
AMD late
nAMD
GA + nAMD
GA pure
Protein Quintile

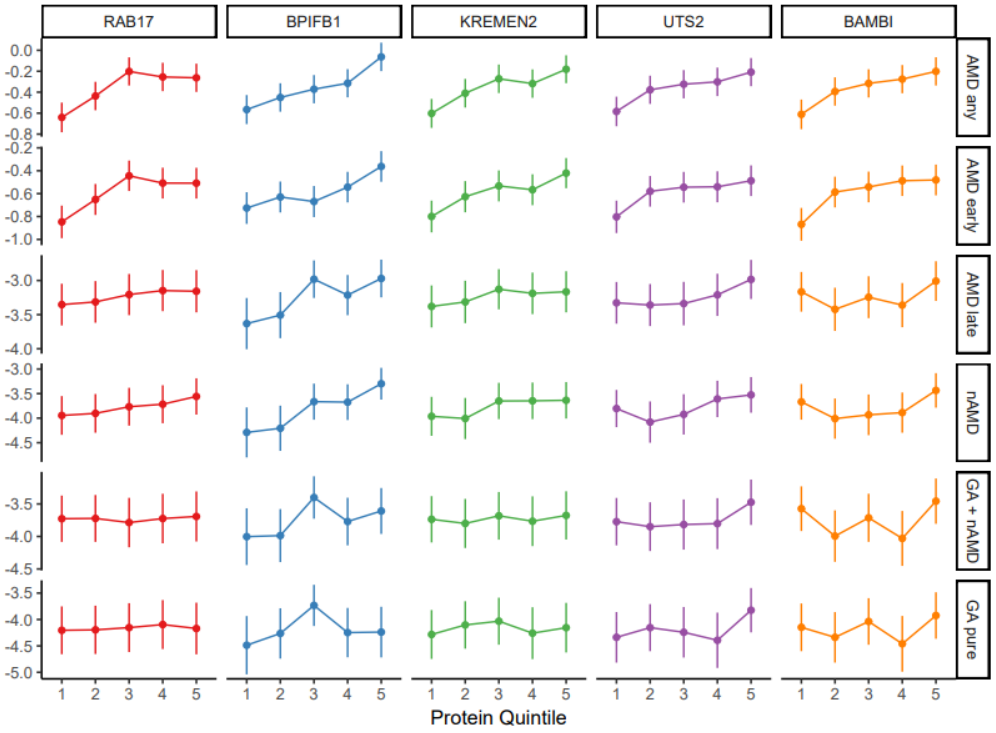
RAB17
BPIFB1
KREMEN2
UTS2
BAMBI
AMD any
AMD early
AMD late
nAMD
GA + nAMD
GA pure
Protein Quintile

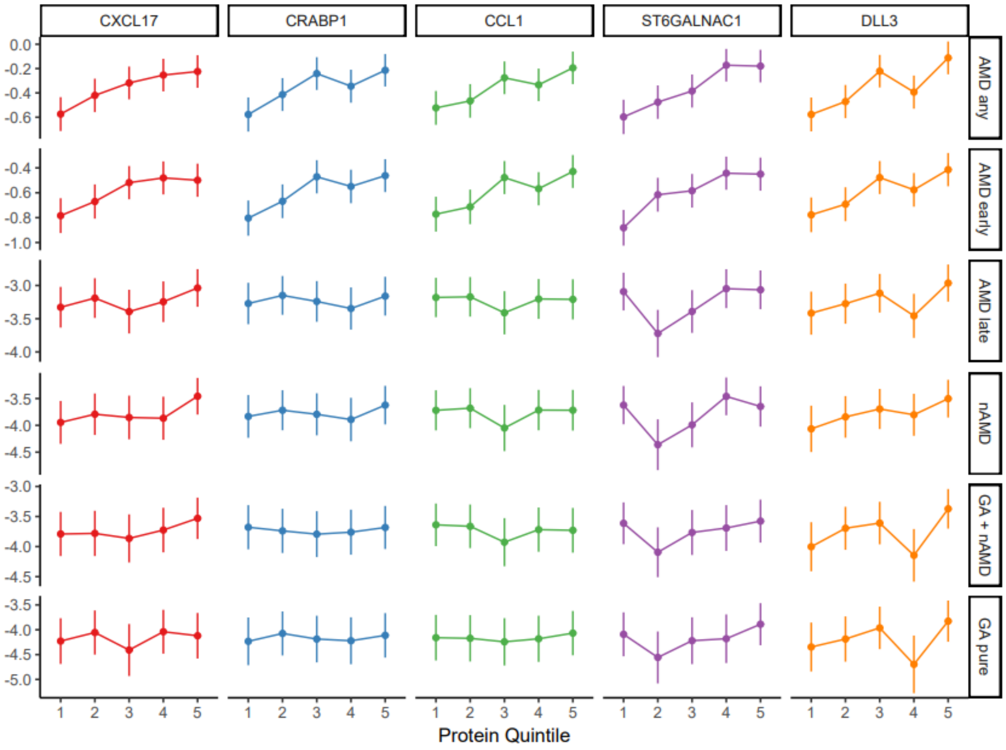
CXCL17
CRABP1
CCL1
ST6GALNAC1
DLL3
AMD any
AMD early
AMD late
nAMD
GA + nAMD
GA pure
Protein Quintile

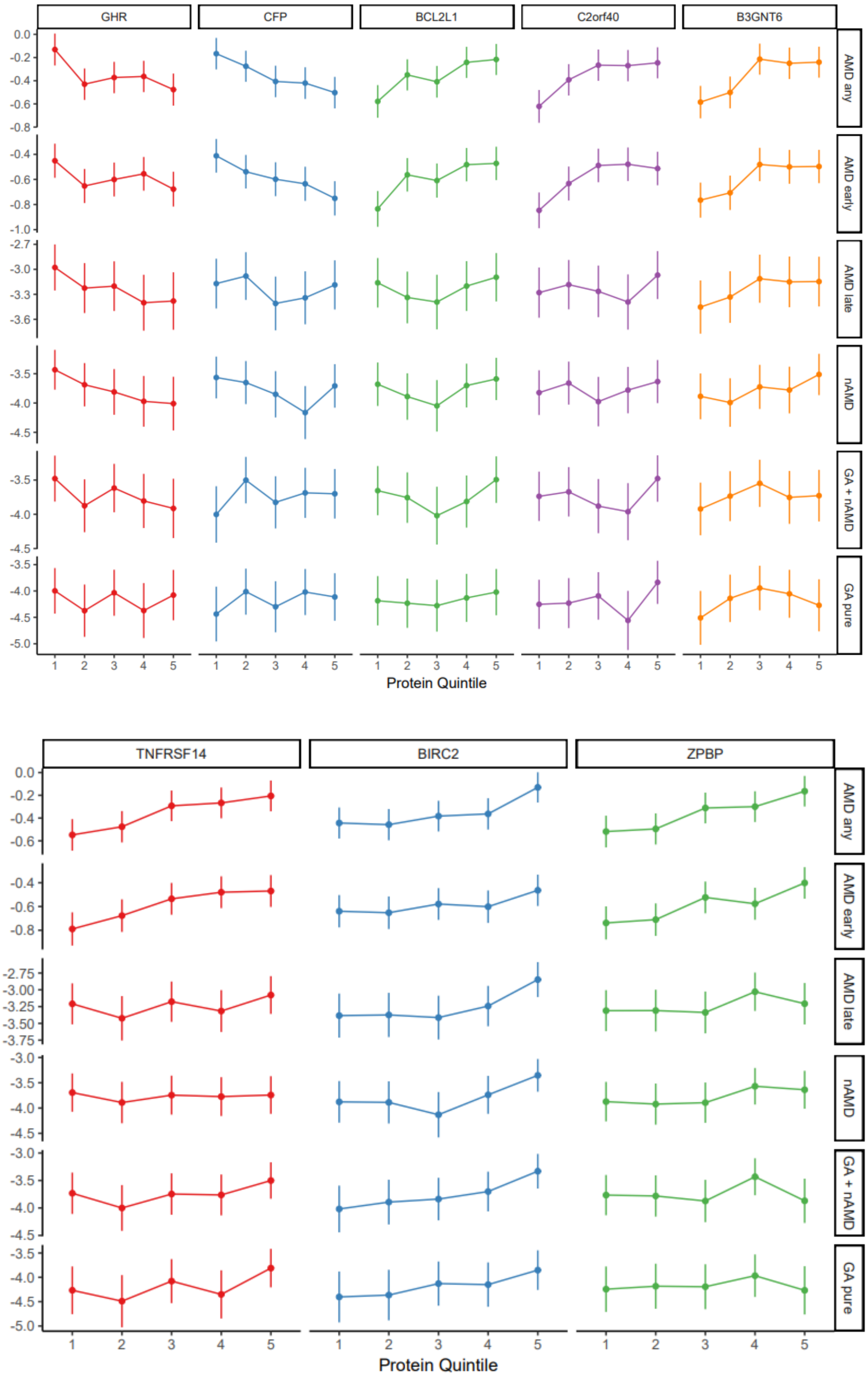


### Supplementary Figure S3

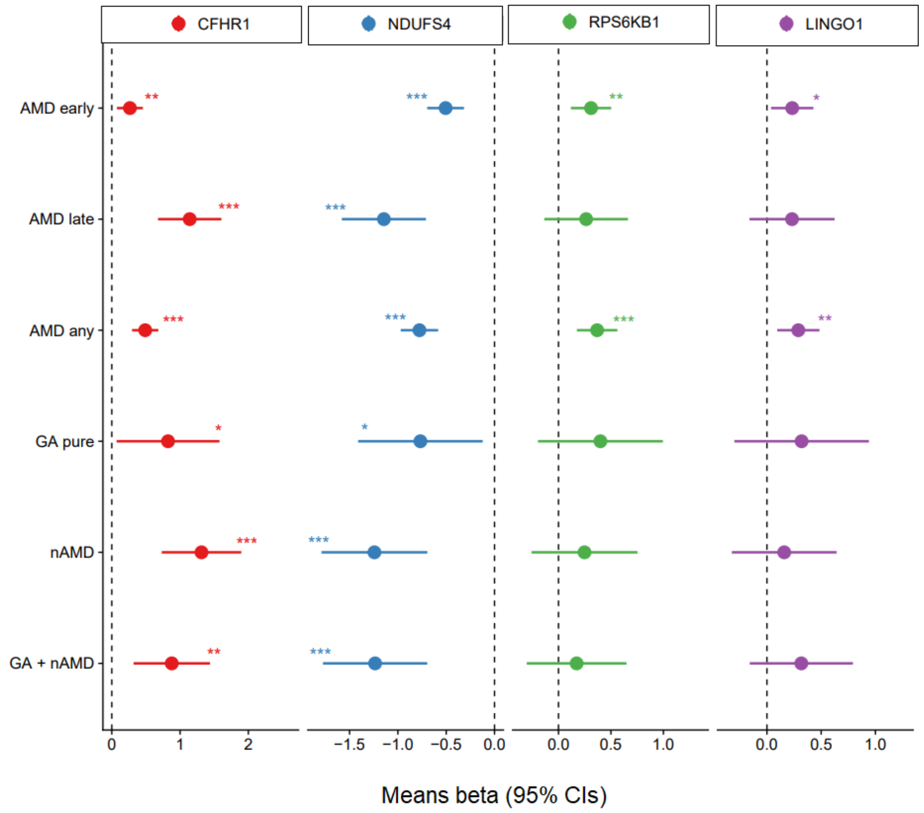
CFHR1
NDUFS4
RPS6KB1
LINGO1
AMD early
AMD late
AMD any
GA pure
nAMD
GA + nAMD
0 1 2
−1.5 −1.0 −0.5 0.0
0.0 0.5 1.0
0.0 0.5 1.0
Means beta (95% CIs)

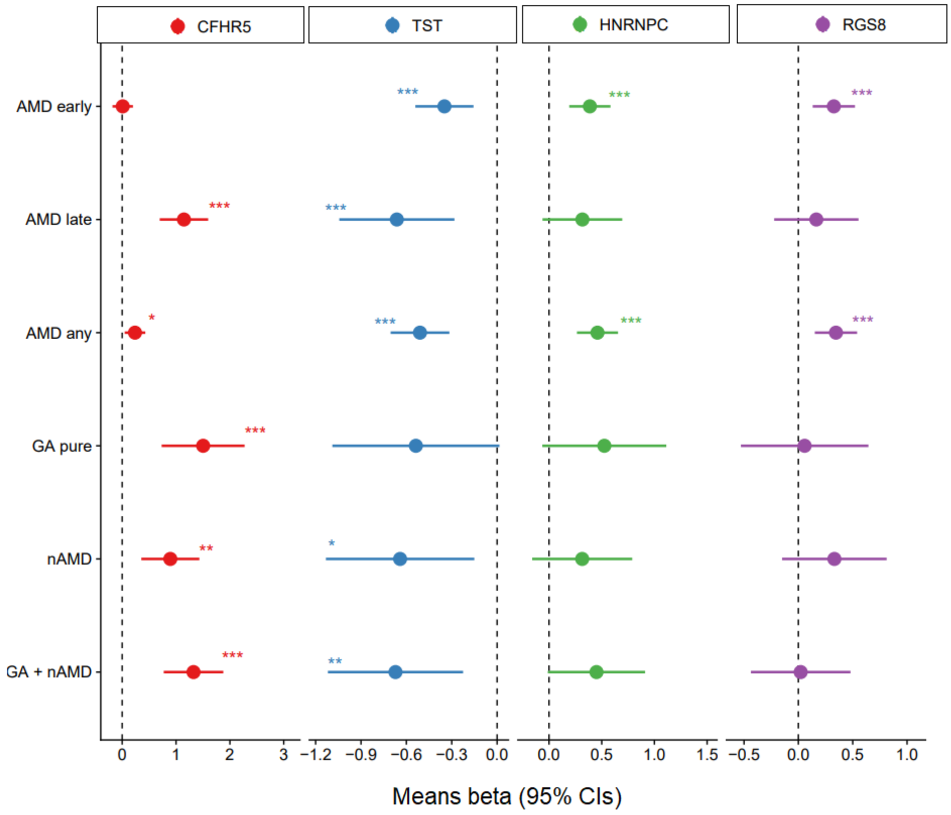
CFHR5
TST
HNRNPC
RGS8
AMD early
AMD late
AMD any
GA pure
nAMD
GA + nAMD
0 1 2 3
−1.2 −0.9 −0.6 −0.3 0.0
0.0 0.5 1.0 1.5
−0.5 0.0 0.5 1.0
Means beta (95% CIs)

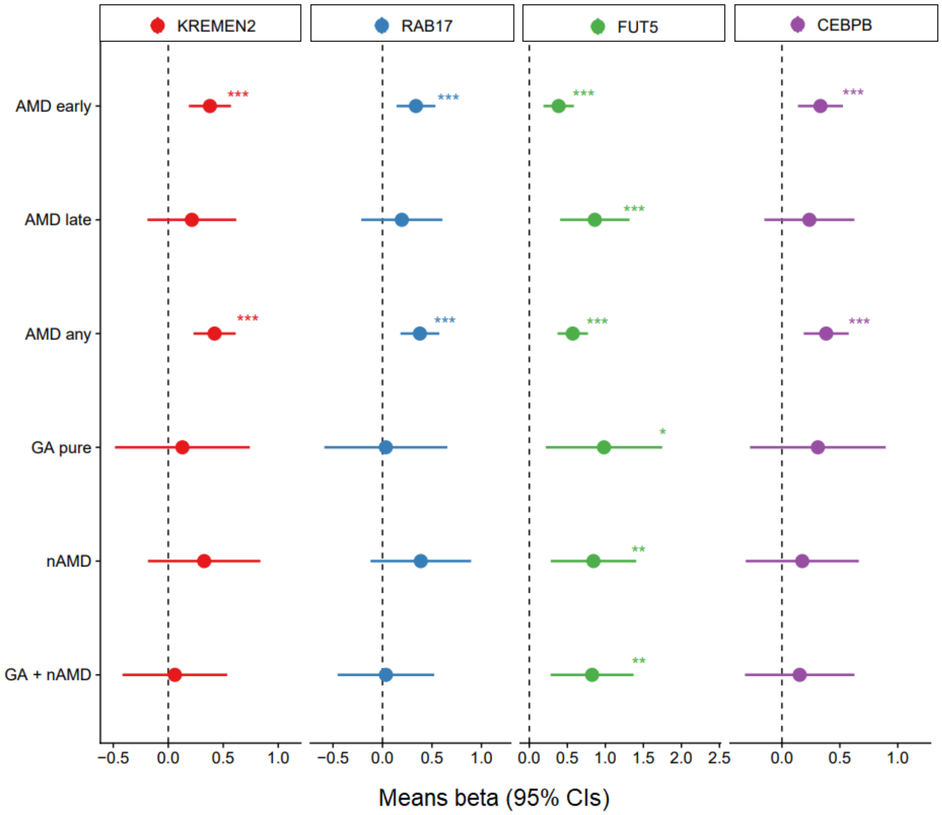
KREMEN2
RAB17
FUT5
CEBPB
AMD early
AMD late
AMD any
GA pure
nAMD
GA + nAMD
Means beta (95% CIs)

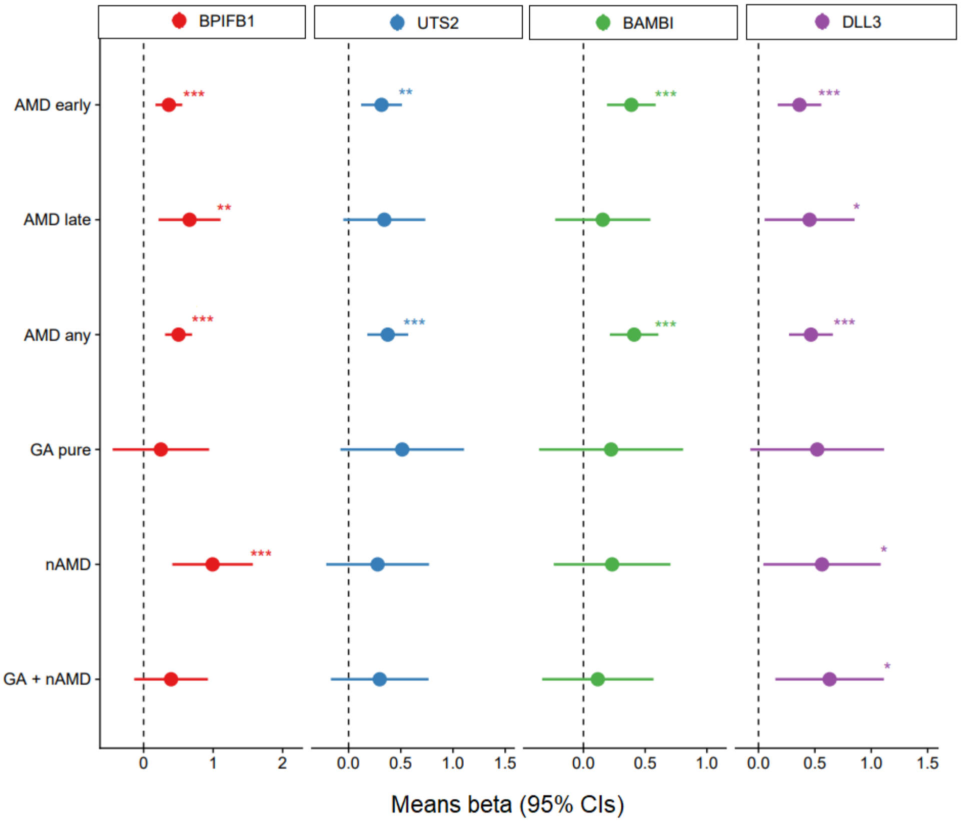
BPIFB1
UTS2
BAMBI
DLL3
AMD early
AMD late
AMD any
GA pure
nAMD
GA + nAMD
Means beta (95% CIs)

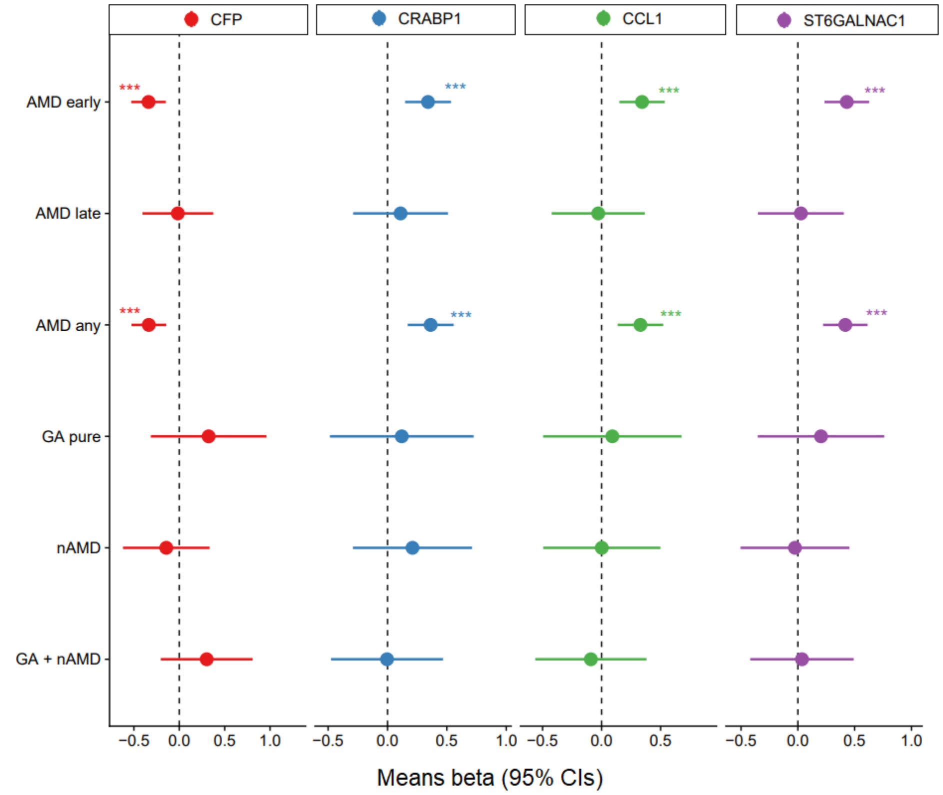
CFP
CRABP1
CCL1
ST6GALNAC1
AMD early
AMD late
AMD any
GA pure
nAMD
GA + nAMD
Means beta (95% CIs)

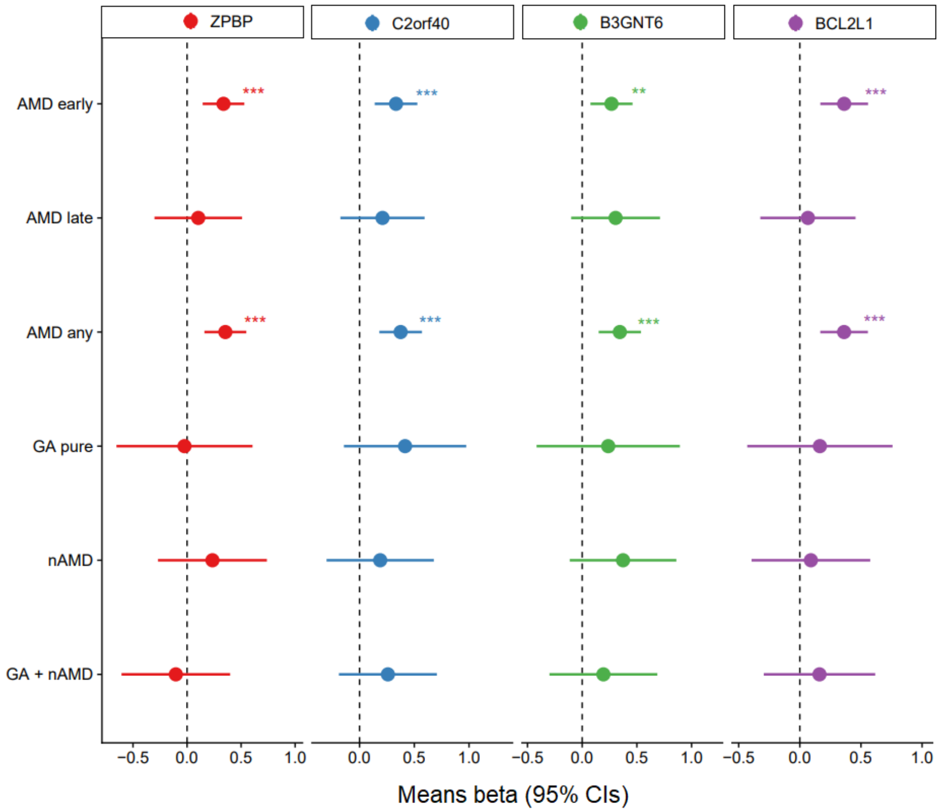
ZPBP
C2orf40
B3GNT6
BCL2L1
AMD early
AMD late
AMD any
GA pure
nAMD
GA + nAMD
Means beta (95% CIs)

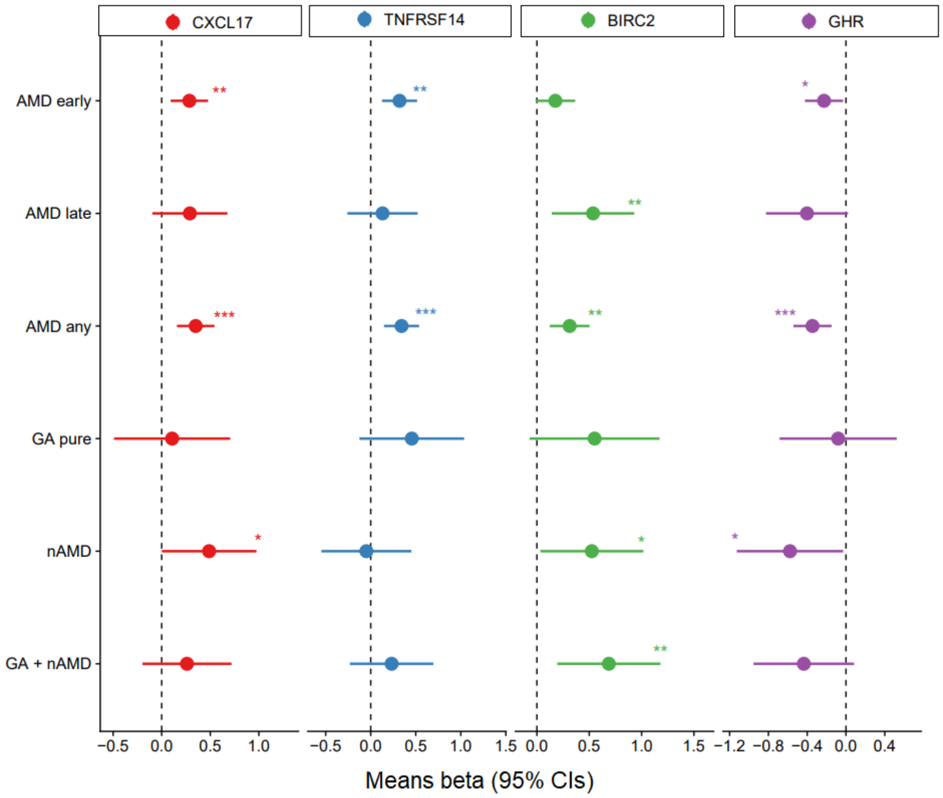


**Supplementary Figure S4**

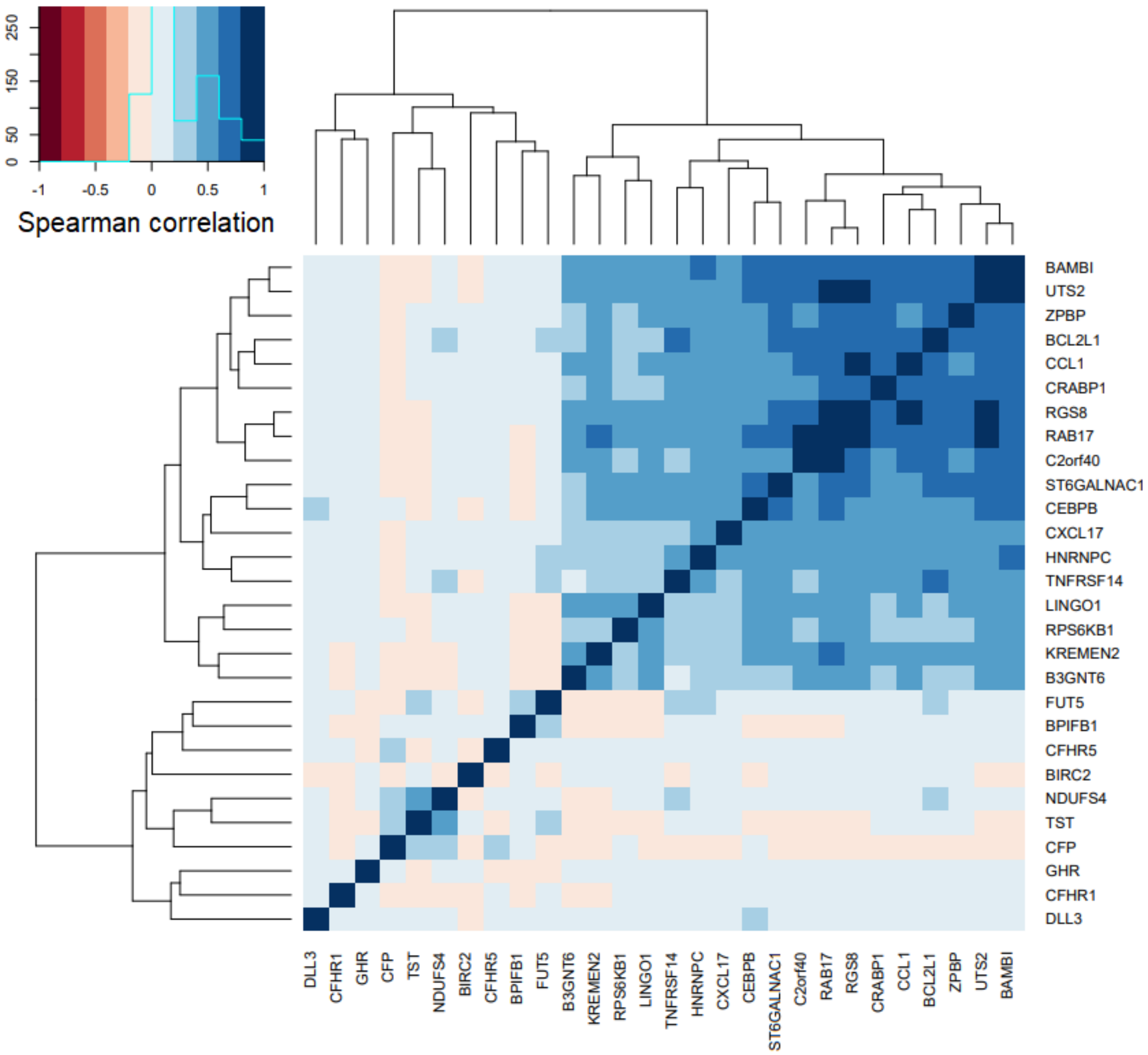


**Supplementary Figure S5**

#### scRNAseq of macular RPE-choroid

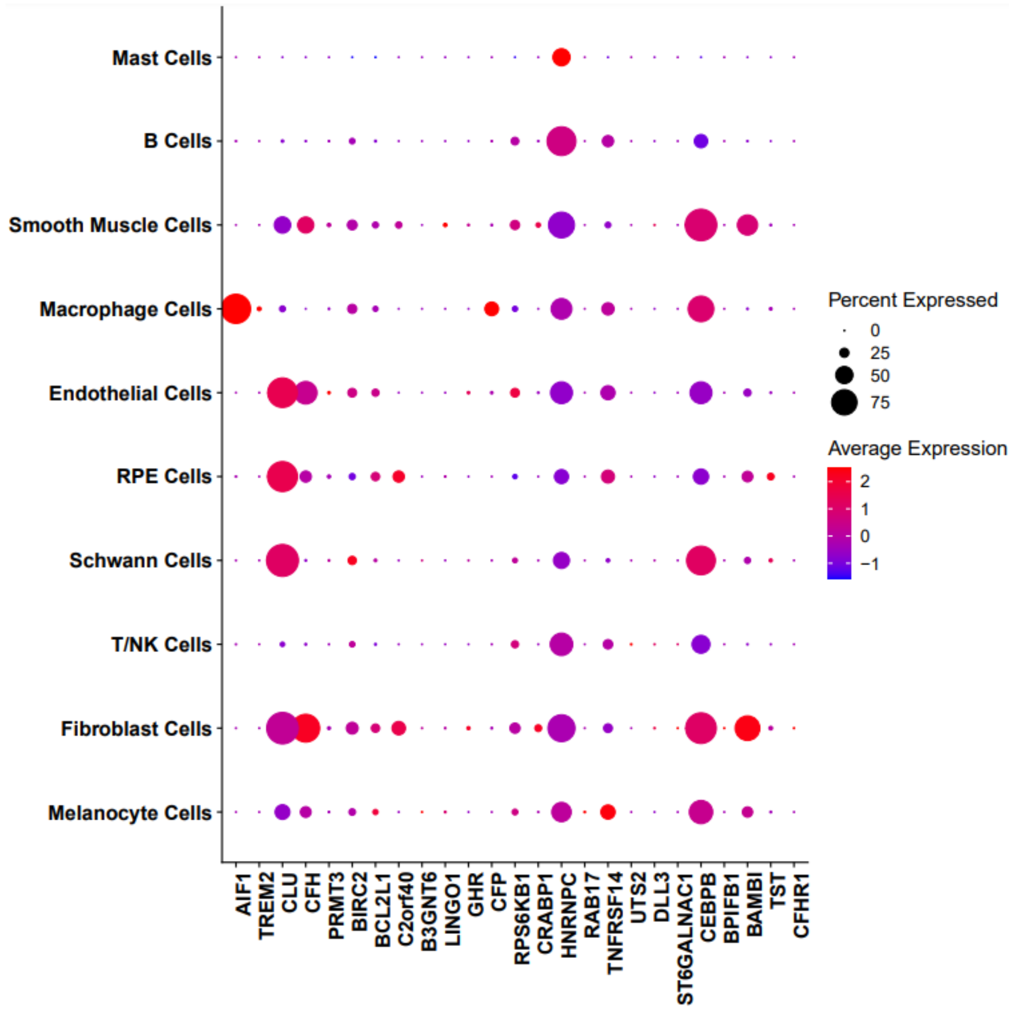


### Supplementary Figure S6

**a**

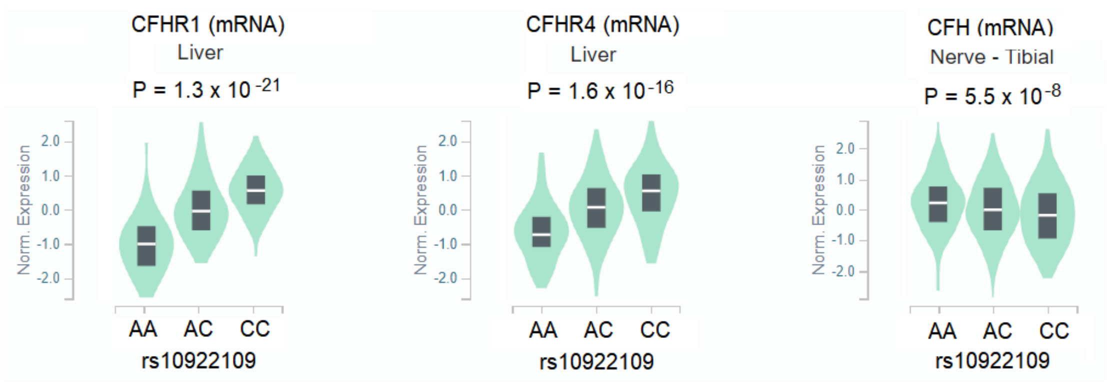


**b**

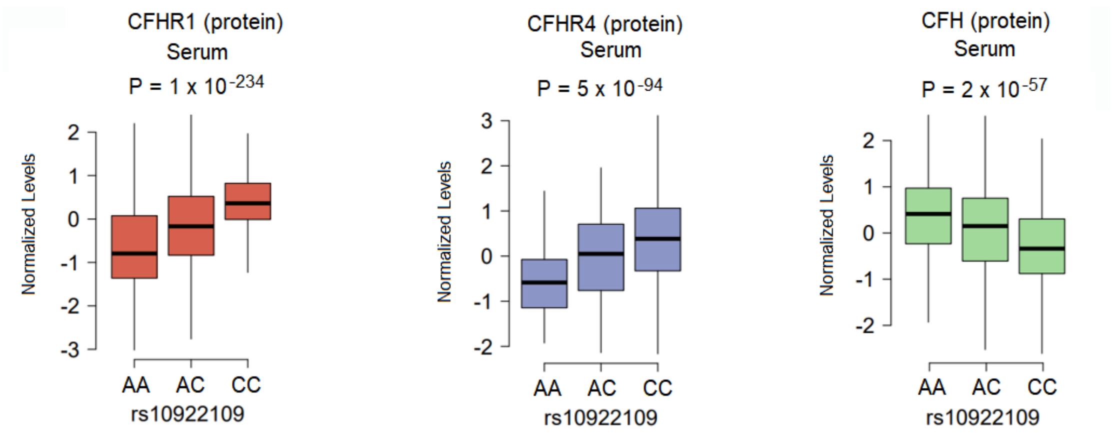


#### Supplementary Figure S7

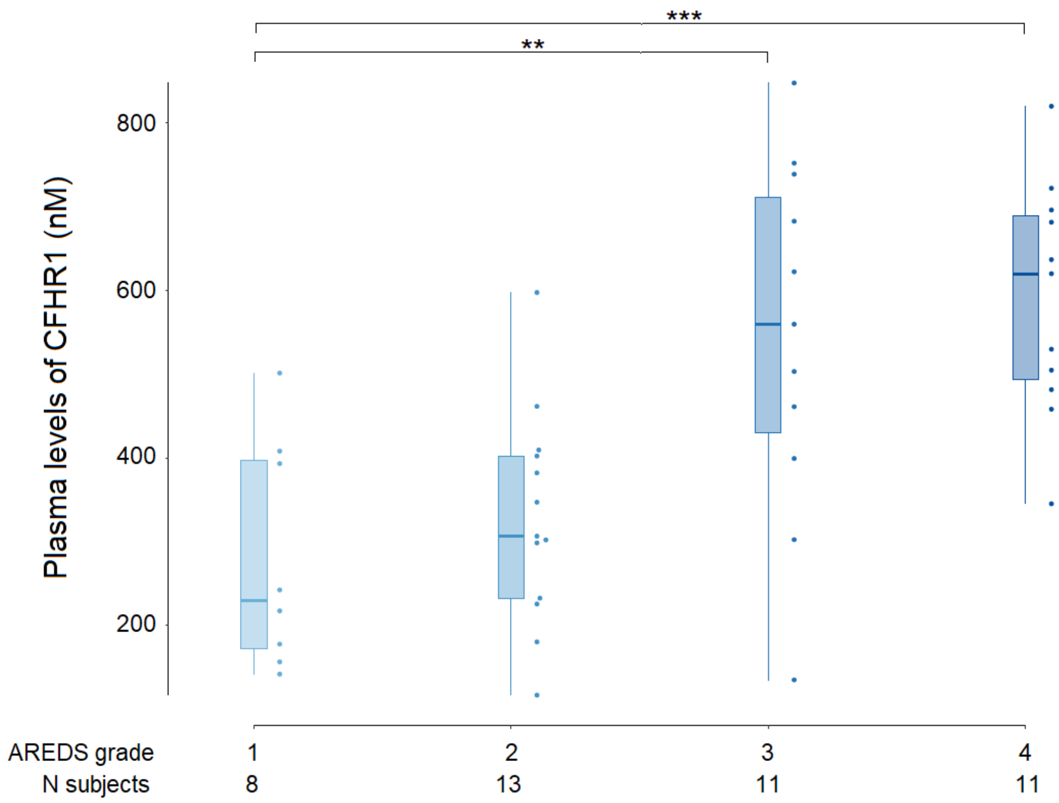


**Supplementary Figure S8**

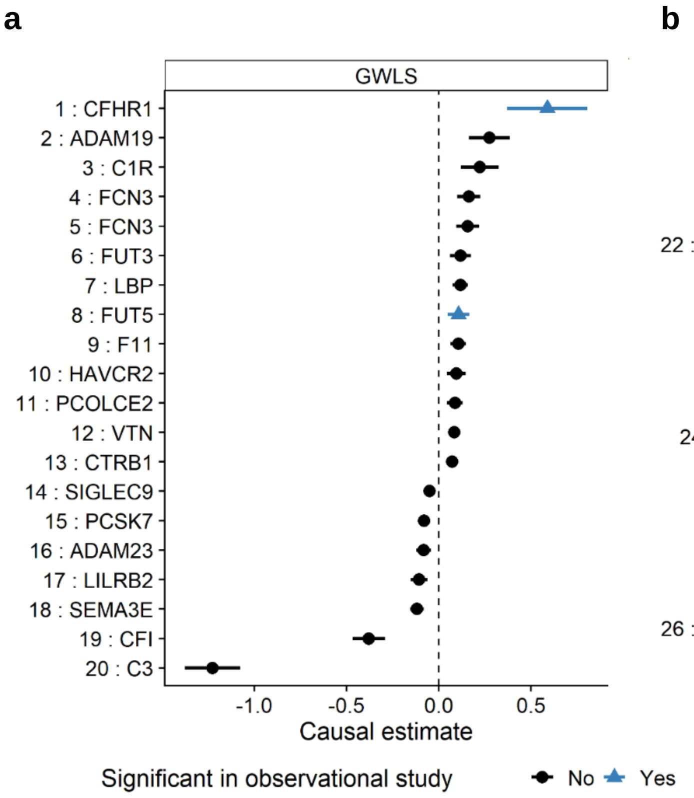


### Supplementary Figure S9
